# SJPedPanel: A pan-cancer gene panel for childhood malignancies

**DOI:** 10.1101/2023.11.27.23299068

**Authors:** Pandurang Kolekar, Vidya Balagopal, Li Dong, Yanling Liu, Scott Foy, Quang Tran, Heather Mulder, Anna LW Huskey, Emily Plyler, Zhikai Liang, Jingqun Ma, Joy Nakitandwe, Jiali Gu, Maria Namwanje, Jamie Maciaszek, Debbie Payne-Turner, Saradhi Mallampati, Lu Wang, John Easton, Jeffery M. Klco, Xiaotu Ma

**Author notes:** correspondence authors, XM; JMK; JE. Contributed equally (Pandurang Kolekar, Vidya Balagopal, Li Dong).

## Abstract

**Background:** Large scale genomics projects have identified driver alterations for most childhood cancers that provide reliable biomarkers for clinical diagnosis and disease monitoring using targeted sequencing. However, there is lack of a comprehensive panel that matches the list of known driver genes. Here we fill this gap by developing SJPedPanel for childhood cancers.

**Results:** SJPedPanel covers 5,275 coding exons of 357 driver genes, 297 introns frequently involved in rearrangements that generate fusion oncoproteins, commonly amplified/deleted regions (e.g., *MYCN* for neuroblastoma, *CDKN2A* and *PAX5* for B-/T-ALL, and *SMARCB1* for AT/RT), and 7,590 polymorphism sites for interrogating tumors with aneuploidy, such as hyperdiploid and hypodiploid B-ALL or 17q gain neuroblastoma. We used driver alterations reported from an established real-time clinical genomics cohort (n=253) to validate this gene panel. Among the 485 pathogenic variants reported, our panel covered 417 variants (86%). For 90 rearrangements responsible for oncogenic fusions, our panel covered 74 events (82%). We re-sequenced 113 previously characterized clinical specimens at an average depth of 2,500X using SJPedPanel and recovered 354 (91%) of the 389 reported pathogenic variants. We then investigated the power of this panel in detecting mutations from specimens with low tumor purity (as low as 0.1%) using cell line-based dilution experiments and discovered that this gene panel enabled us to detect ∼80% variants with allele fraction of 0.2%, while the detection rate decreases to ∼50% when the allele fraction is 0.1%. We finally demonstrate its utility in disease monitoring on clinical specimens collected from AML patients in morphologic remission.

**Conclusions:** SJPedPanel enables the detection of clinically relevant genetic alterations including rearrangements responsible for subtype-defining fusions for childhood cancers by targeted sequencing of ∼0.15% of human genome. It will enhance the analysis of specimens with low tumor burdens for cancer monitoring and early detection.

## Background

Extensive insights on the genetic underpinnings (i.e., driver alterations) of childhood cancers ^1–3^ have been uncovered in the past decade using next generation sequencing. To date, diagnostic sequencing has become part of clinical service in some institutions^4–6^. Although whole genome, and to lesser extent whole exome sequencing, is preferrable to maximize the detection of cancer-associated variants in the clinical setting, there are significant resource and infrastructure requirements for these modalities that are not amenable to the majority of clinical labs. Further, the broad coverage of whole genome and whole exome sequencing renders it challenging to achieve ultra-deep sequencing that is essential for the analysis of specimens with low tumor purity such as for detecting minimal residual disease and for disease monitoring. Targeted gene panel-based sequencing holds the promise to address these challenges.

Although there have been multiple gene panels designed for adult cancers, such as MSK-IMPACT^6^, currently there is no comprehensive gene panel for pediatric cancers. This is important considering the recent pan-cancer study of 1,699 childhood cancers that demonstrated a dramatic difference between adult and childhood cancers, where 55% of the 142 driver genes in pediatric cancers are not found in adult pan-cancer studies ^3^. In this study, 62% of driver alterations in childhood cancers are copy number alterations (CNVs) or structural variations (SVs) whose boundaries typically do not fall into protein coding regions. Indeed, our recent study of oncogenic fusions^7^ indicated that 55.7%, 22.5%, and 18.5% of pediatric leukemia, brain, and solid tumors have subtypes defined by oncogenic fusions, for which the DNA breakpoints typically fall into intronic regions. These facts render base pair level ascertainment of driver alterations in childhood cancers challenging by using conventional capture sequencing kits such as exome sequencing and call for a dedicated gene panel for pediatric cancers that includes both coding and non-coding targets to maximize the detection of key alterations in pediatric cancers.

Here, we highlight the prominent features of our pan-cancer gene panel (termed SJPedPanel) for childhood cancers by comparison with five existing cancer gene panels. We validate its superior coverage of genes relevant to childhood cancers using a well-described real-time clinical cohort via *in silico* analysis, followed by re-sequencing a subset of these cases for experimental validation. We also demonstrate the power of our gene panel in detecting rare variants using ultra-deep sequencing via serial dilution experiments, as well as disease monitoring in remission samples from acute myeloid leukemia (AML) patients.

## Results

### Panel design

We designed our SJPedPanel by integrating findings from 44 published tumor-normal paired genomics studies of childhood cancers that spans leukemia, brain, and solid tumor ^2–4,8–48^. SJPedPanel includes 1.069 million exonic base pairs from 5,275 coding exons for detecting protein coding mutations in 357 known driver genes for childhood cancers [**Fig. 1a**; see Additional file 1: Table ST1, and Additional file 2 for plots of chromosomal distribution of genes generated using an R/Bioconductor package chromPlot ^49^]. To account for the structural variations (SVs) that result in subtype-defining oncogenic fusions for which DNA breakpoints typically fall in intronic regions ^7^, 1.438 million base pairs from 297 introns of 94 genes [Additional file 1: Table ST1] were included. Moreover, 0.209 million bases from promoter regions were targeted for detecting promoter alterations including rearrangements and point mutations such as *TAL1* in T-ALL^50^ [**Fig. 1b**]. Highly recurrent oncogenes (*MYCN* in neuroblastoma^3^) and tumor suppressor genes (such as *CDKN2A*, *PAX5*, and *SMARCB1*) were targeted by probes tiling the entire gene region for detecting CNVs. To account for the fact that breakpoints of structural alterations can fall outside gene regions, we extended the target regions to frequent DNA breakpoints by using patient data from ProteinPaint ^51^ and GenomePaint ^52^. Collectively, 2.82 million base pairs were designed for potential SNV, Indel, SV and CNV/LOH driver alterations. Notably, a few known childhood cancer drivers are intentionally excluded due to genomic space considerations. For example, *MECOM* ^53^ and *GFI1B* ^54^ are known to be involved in enhancer-hijacking alterations and were excluded due to the large space needed to cover the many possible breakpoints.

**Fig. 1.**
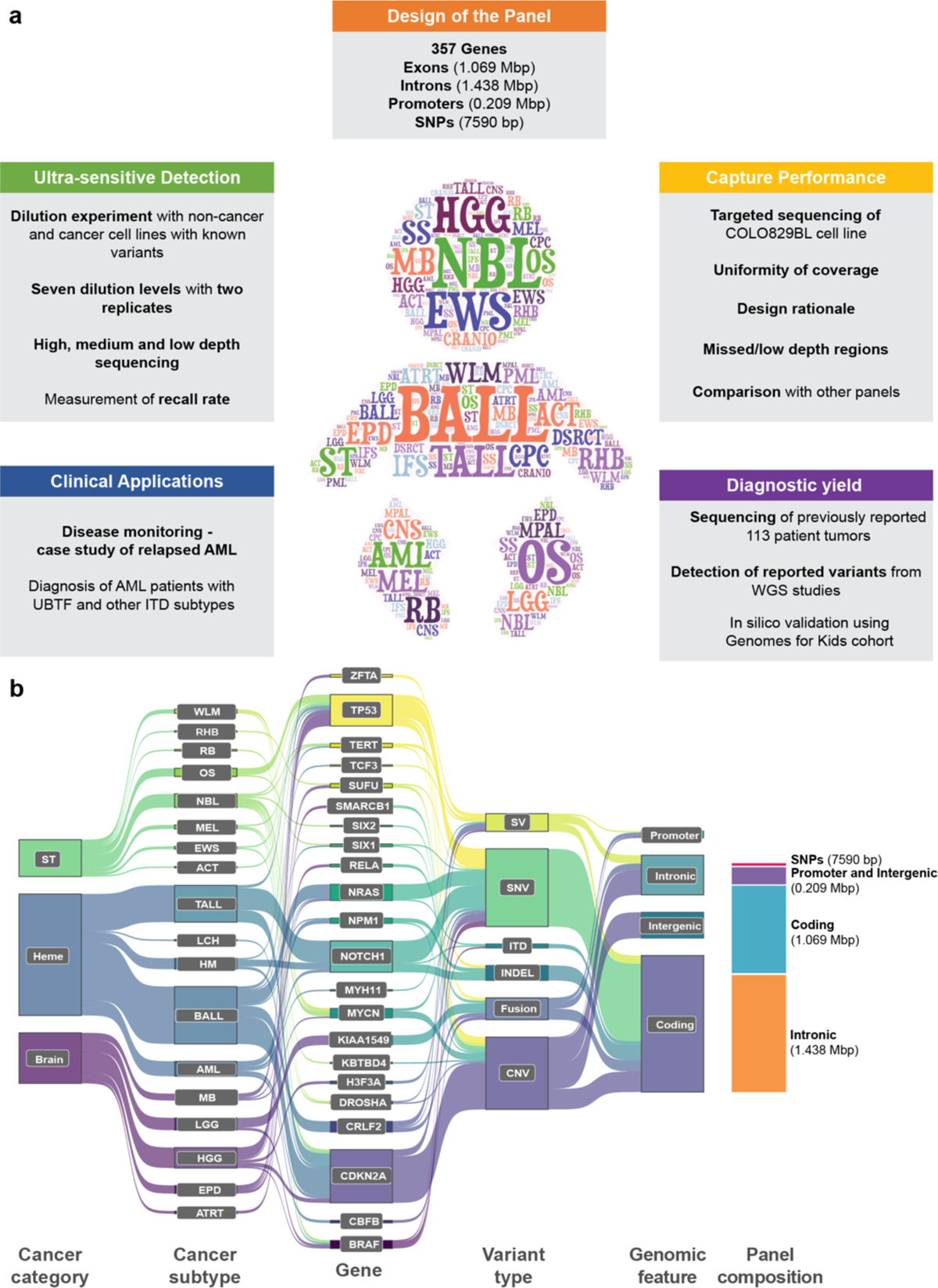
Design of pediatric cancer gene panel. (a) This study includes panel content, investigation of ultra-sensitive detection, capture performance, diagnostic yield, and clinical applications. (b) A Sankey diagram ^55^ showing spectrum of childhood cancers (Heme: hematological malignancies; ST: solid tumors; Brain: brain tumors), cancer subtypes, genes, variant types, and genomic features targeted by SJPedPanel. Stacked bar plot at the right end shows space distribution of different genomic features covered by SJPedPanel.

In addition, 7,590 SNPs were selected for detecting copy number variations and loss of heterozygosity (CNV/LOH) across the genome [see Additional file 1: Table ST2]. The median distance between these SNPs is 332 Kb, with 25^th^ and 75^th^ quantile being 60 Kb and 593 Kb, respectively [see Additional file 3: Fig. S1a]. Notably, >80% of these SNPs have population frequency between 40% and 60% [see Additional file 3: Fig. S1b], which ensures that that nearly 50% of patients are heterozygous at each SNP site. Therefore, around 3,000 (=7590×0.5×0.8) heterozygous SNPs are expected for each patient, which leads to a theoretical resolution of ∼1 Mb for CNV/LOH detection. The number of SNPs chosen per chromosome is roughly proportional to the lengths of chromosomes [see Additional file 3: Figs. S1c-d, and Additional file 4 for plots of chromosomal distribution of SNPs generated using an R/Bioconductor package IdeoViz ^56^]. Considering the read length and the insert length (for target capture and sequencing), these 7,590 SNPs actually occupy ∼250*7590=1.8975 million base pairs. Thus, our panel consisted of ∼4.7 million base pairs, or ∼0.15% of the human genome. The compact size of our panel enables us to reach 30,000X at the cost of a standard WGS (∼30X) per sample, thus enabling cost-effective cancer monitoring and/or early detection [**Fig. 1a**]. The gene panel was manufactured by Twist Bioscience.

### Comparison of gene content with other panels

We first compared the gene content between our panel and five other commonly used commercial panels for childhood cancers, including FoundationOne Heme, FoundationOne CDx ^57,58^, MSK-IMPACT ^6,59^, OncoKids ^5^, and Oncomine Comprehensive assay v3 (OCAv3) ^60^ [Additional file 1: Tables ST3a and ST3b]. We used the list of 183 driver genes reported in two recent childhood pan-cancer studies ^2,3^ involving 2,578 cases. As seen in **Table 1**, SJPedPanel covers 159 (87%) genes whereas all other panels covered <60% of the reported pediatric cancer driver genes [**Fig. 2a** and Additional file 1: Table ST4].

**Fig. 2.**
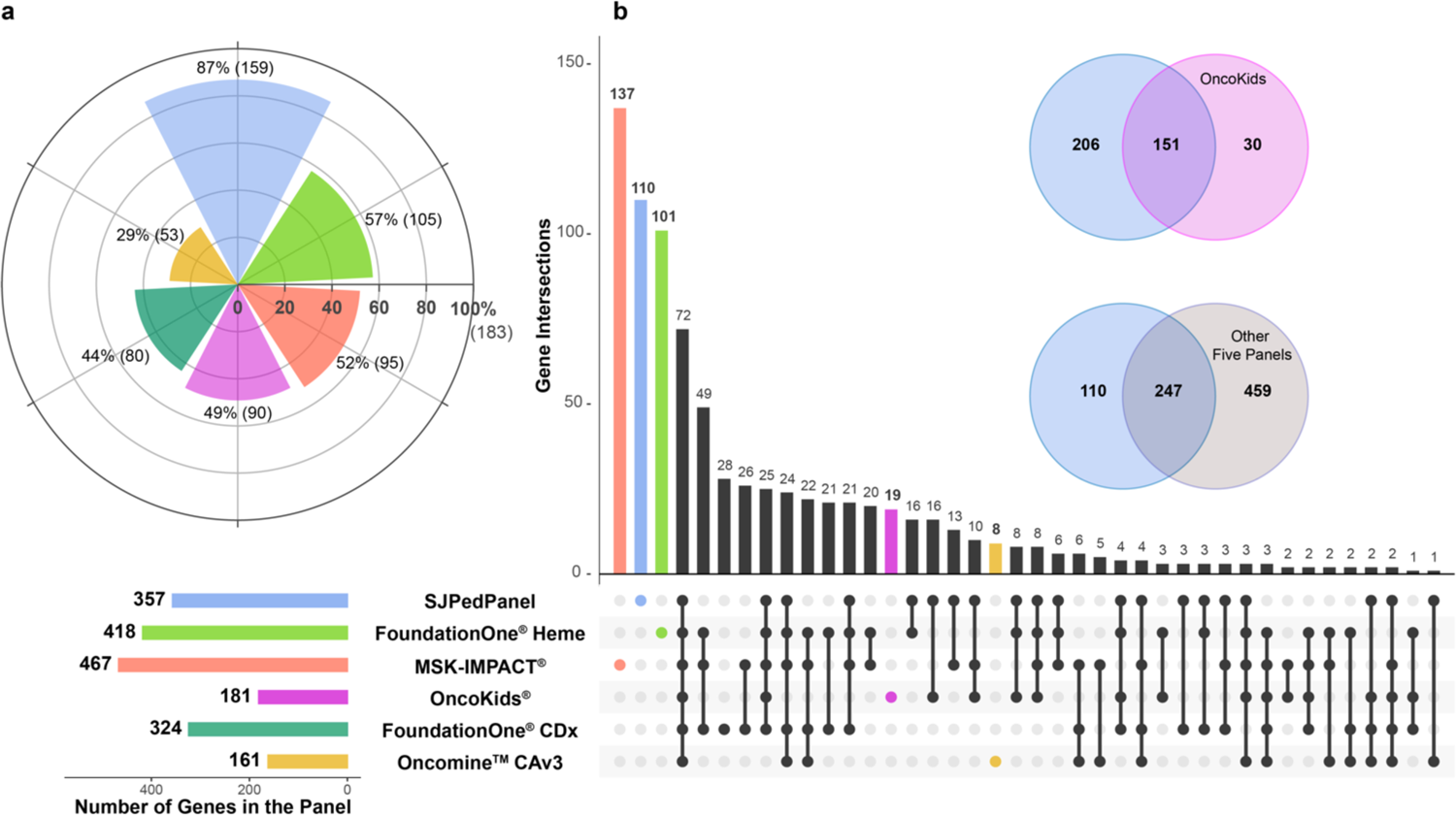
Comparison of gene content between SJPedPanel with other panels. **(a)** Pediatric cancer relevance of 6 panels based on coverage of 183 driver genes from childhood pan-cancer studies ^2,3^. The horizontal bars at the bottom indicate numbers of genes designed in each panel coded by corresponding color. **(b)** Analysis of common and unique genes among 6 panels using UpSet plot ^68^. Venn diagrams indicate intersection of SJPedPanel genes with OncoKids, and other fiver panels combined.

**Table 1.** Overview of the panels selected for comparison.

| Panel name | Focus area | No. of genes | Genes screened for |  |  | SNPs | %Pediatric cancer driver genes covered out of reported 183 genes<sup>†</sup> | Publication |
| --- | --- | --- | --- | --- | --- | --- | --- | --- |
|  |  |  | Coding exons | Selected introns | Promoter regions |  |  |  |
| SJPedPanel | Pediatric cancers | 357 | 357 | 94 | 10 | 7590 | 87% | This study |
| FoundationOne® Heme | Adult blood cancers | 418 | 408 | 31 |  |  | 57% | He et al., 2016<sup>66</sup> |
| MSK-IMPACT® | Adult solid tumors | 468<sup>‡</sup> | 468 | 13 | 1 | 862 | 52% | Cheng et al., 2015<sup>6</sup> |
| OncoKids® | Pediatric cancers | 181 | 137 | 44 | 1 |  | 49% | Hiemenz et al., 2018<sup>5</sup> |
| FoundationOne® CDx | Adult solid tumors | 324 | 309 | 34 | 1 |  | 44% | Whitepaper by Company<sup>§ 58</sup> |
| Oncomine™ Comprehensive Assay v3 | Adult cancers | 161 | 146 | 15 | 1 |  | 29% | Hovelson et al., 2015<sup>67</sup> |

<sup>†</sup>: 183 pediatric cancer genes are reported in two pediatric pan-cancer studies, Ma et al., 2018<sup>3</sup> and Grobner et al., 2018<sup>2</sup>
<sup>‡</sup>: MSK-IMPACT panel has been reported to consist of 468 genes considering 2 different transcript isoforms for CDKN2A gene, however based on unique gene names MSK-IMPACT panel consists of 467 genes.
<sup>§</sup>: Sources of content for all the panels are available in Additional file 1: Table ST3a

A comparison of gene names among the panels indicated that SJPedPanel has unique coverage of 110 genes [**Fig. 2b**, Additional file 1: Table ST3b] when compared to the other panels combined, such as *DGCR8* and *SIX1* for Wilms tumor ^61^, *SHH* for medulloblastoma ^62^, *ZFTA* for ependymoma ^22,63^, *UBTF*-TD ^64^ and *PICALM* ^64,65^ for AML. Among all the panels, SJPedPanel provides the largest intronic regions (297 introns from 94 genes; **Table 1**) responsible for rearrangements that generate fusion oncoproteins.

On the other hand, among the 459 genes specific to other panels, only three genes were reported in two recent pediatric pan-cancer studies with low patient frequencies (*ZNF217: 0.59%, PCBP1: 0.31%, and CARD11: 0.21%*) ^2,3^. MSK-IMPACT panel has the maximum number of genes (467), of which 137 are exclusive from other panels ^6,59^. Most of these genes are relevant to adult cancers with the highest concentration in adult solid tumors ^6,59^. Similarly, FoundationOne Heme panel consists of 418 genes with a focus on adult hematological malignancies ^57,66^. OncoKids is the only other pediatric cancer panel under comparison and covers 30 genes that are not included in our panel [see **Fig. 2b** and Additional file 1: Table ST3b]. *MECOM* is excluded due to large space needed for the diverse promoter hijacking events ^53^.

*CALR, RARA* and *SS18* were not included due to an overall paucity of alterations in these genes in current pediatric genome cohorts. Collectively, SJPedPanel offers by far the most comprehensive coverage of genetic alterations for the study of childhood malignancies.

### Capture performance of the panel

A critical consideration in genomic sequencing (especially in panel sequencing) is the coverage uniformity. To study this question, we sequenced four targeted sequencing libraries (C1-C4) prepared using COLO829BL (ATCC #CRL-1980), a non-cancer cell line that has been extensively used in literature for clinical proficiency testing or benchmarking ^69,70^. To ensure reproducibility, technical replicates were generated to achieve high (C1, C2, ∼2000X) and low depth (C3, C4, ∼200X) of sequencing. Libraries in each set were sequenced with either the Illumina NovaSeq 6000 (C1, C3) or NextSeq 500 (C2, C4).

As expected, the average depth was highly correlated (*r^2^*: 0.98) with the number of raw reads sequenced [Additional file 3: Supplementary Fig. S2a]. With this data, we investigated the capture uniformity at base pair level [**Fig. 3a-b**] and at region level [**Fig. 3c-d**]. Because highly uniform capture data would ensure most bases/regions to have similar depth (therefore a histogram with very small standard deviation), we choose to use coefficient of variation (*C_V_*, defined by *σ / μ* of the histogram) to measure sequencing uniformity. Here *σ* and *μ* are the estimate of standard deviation and mean, respectively, by trimming 2.5% of extreme values from both ends of the histograms [**Fig. 3**]. At base pair level, we observed that *C_V_* is close to 0.35 for libraries sequenced by NovaSeq, and to be between 0.37 and 0.38 for libraries sequenced by NextSeq. At region level, NovaSeq data has *C_V_* close to 0.25, while NextSeq data has *Cv* range from 0.22 to 0.28.

**Fig. 3.**
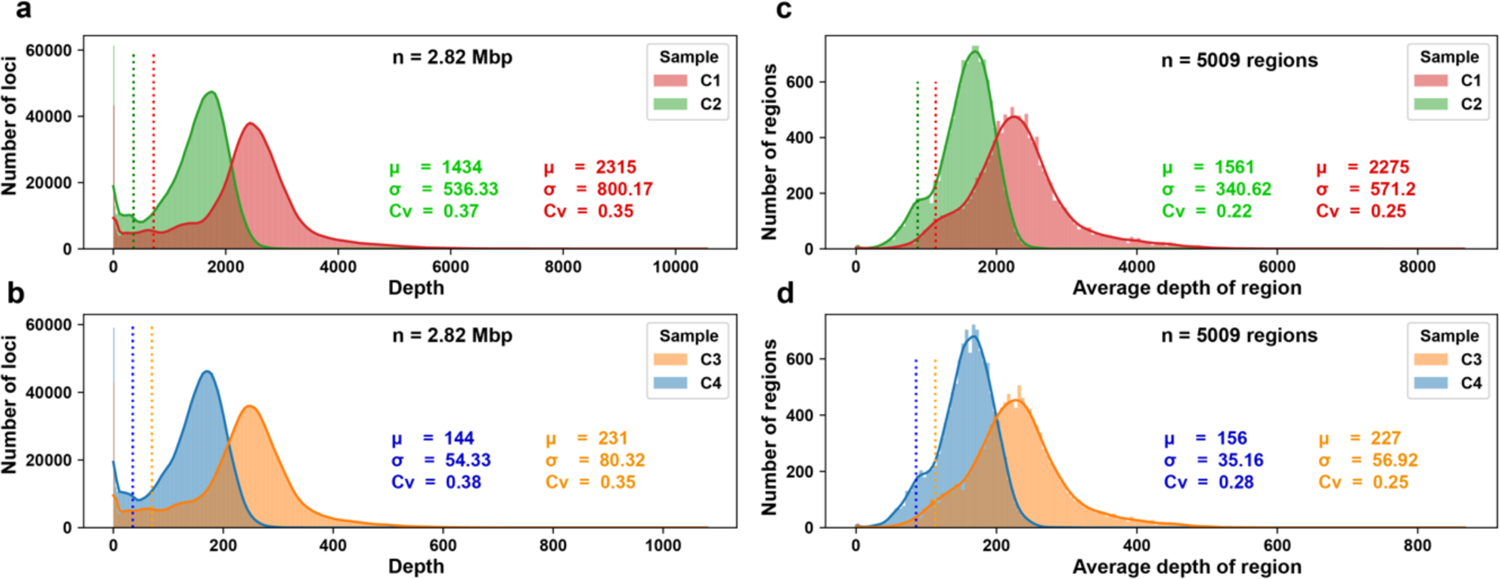
Capture uniformity per base (a, b) and per region (c, d) of the panel. Uniformity of coverage across in the SJPedPanel for high depth samples sequenced on Illumina NovaSeq 6000 (C1) and NextSeq 500 (C2) respectively; and low depth samples sequenced on Illumina NovaSeq 6000 (C3) and NextSeq 500 (C4) respectively. The histograms are made at base pair level (2.82 Mbp) **(a, b)** and region level (n=5009) **(c, d)**. The vertical dotted lines indicate (*μ - 2σ*) of the respective distributions. The statistical parameters (*μ*: average depth, *σ*: standard deviation, *Cv*: coefficient of variance) were calculated by trimming observations in upper and lower 2.5 percentiles. All the sample and region level QC parameters are available in the Additional file 1: Tables ST5 and ST6.

Overall, the standard deviation is less than or around 1/3 of the mean, which ensures that most of the target bases/regions are sufficiently covered. Using the two-sigma rule (that approximates the 95% confidence interval), we also measured the percentage of bases/regions with depth higher than (*μ - 2σ)*. We found that 97% and 95% of bases have depth higher than this threshold for NovaSeq data and NextSeq data, respectively [Additional file 1: Table ST5 and Additional file 3: Fig. S2b]. Similar trends were observed from the region level analyses [**Figs. 3c-d** and Additional file 1: Table ST6]. These data indicated that SJPedPanel has satisfactory capture efficiency that is reproducible over different sequencing platforms.

We next analyzed regions that are consistently poorly covered (i.e., less than *μ - 3σ*) in the COLO829 data. We identified 27 regions (26 regions are small exons), of which 10 regions consistently showed no coverage across all the four samples [see column “Remark” in Additional file 1: Table ST6]. These 27 regions occupy 8091 bp (∼0.3%) of the panel and more than half of these bases (4972 bp) belong to only two regions, *NUTM2A* (3152 bp including intron 1 with 2050 bp) and *DUX4* (1820 bp including exon 1) [see Additional file 1: Table ST7 and Additional file 3: Supplementary Fig. S3]. For *STAG2* the coverage is slightly below the pre-defined cut-offs for 3 regions (351 bp) (e.g. ∼ 500X for C1 sample, Additional file 1: Table ST7). While looking for the potential reasons for poor coverage of these regions, we observed that flanking regions (+/- 50 bp) of most of these poorly covered regions comprise either high GC content, such as exon 1 of *MLLT1* (94% GC), or homopolymer runs, such as T-runs around three regions of *STAG2* gene [see Additional file 1: Table ST7], which can be informative for future optimization.

Out of the five panels compared, only MSK-IMPACT panel was reported to have 31 consistently poorly covered regions ^59^, which all happened to be targeted by SJPedPanel as well. Interestingly, SJPedPanel demonstrated sufficient depth of coverage (> (*μ - 2σ)* of respective COLO829BL sample level cut-offs) for 29 out of the 31 regions [Additional file 1: Table ST8]. The remaining two regions, exon 2 of *NOTCH2* and exon 15 of *PMS2,* consistently showed poor coverage as in MSK-IMPACT panel and were also part of the 27 poorly covered regions of SJPedPanel discussed above [Additional file 1: Table ST7].

Similarly, we analyzed the depth of coverage at designed SNPs. Notably, 99.5% of all the 7590 SNPs have depth more than (*μ - 2σ*) of the respective sample level cut-offs [Additional file 1: Table ST9; see Additional file 3: Supplementary Fig. S4a for median and minimum depth of coverages for SNPs]. As expected, the variant allele frequencies (VAF) of all the SNPs in control COLO829BL samples were clustered around either 0, 0.5 or 1 [Additional file 3: Supplementary Fig. S4b]. A total of 3300 heterozygous SNPs (0.3 σ; VAF σ; 0.7) are observed in COLO829BL, supporting the informativeness of our designed SNPs as mentioned above (∼3000 heterozygous SNPs expected from any donors).

### *In silico* comparison of SJPedPanel and WES in a real-time clinical genomics cohort

We next compared the coverage of reported pediatric cancer alterations using whole exome sequencing (WES), because WES is effectively a capture-sequencing kit that targets coding exons of all genes rather than a panel of genes (thus an upper bound of all coding-region-based gene panels). We first asked whether our panel could offer comparable coverage of driver alterations (with a focus on coding SNVs and Indels) in pediatric cancers to WES. The recently published “Genome for Kids” (G4K) study ^71^ reported pathogenic and likely pathogenic (P/LP) variants (called driver alterations hereafter) using three platform sequencing (WGS, WES and RNAseq) from 253 pediatric cases that encompassed 20 cancer subtypes in a real-time clinical genomics setting, thus enabling us to assess the potential of SJPedPanel to cover driver alterations from diverse childhood cancer types. Here, we performed *in silico* analysis of regions targeted by SJPedPanel and WES using the curated positions of 485 driver alterations (including SNV/Indel/SV/ITD; Additional file 3: Supplementary Fig. S5a) from the G4K study [see Additional file 1: Table ST10].

SJPedPanel covered 86% of the 485 reported driver alterations as compared to 76% by WES [**Fig. 4a**, last pair of bars for “All” variants with gray background]. We next classified the variants into SNV, Indel, Fusion/SV (structural rearrangements that result in fusion oncoproteins), Other SV (structural rearrangements that do not result in fusion oncoproteins but affect cancer driver genes such as disrupting tumor suppressor genes), and ITD, by using the class labels in the G4K study ^71^. As expected, WES only covered 12% of the Fusions/SV that have either of the breakpoints in exonic regions, while SJPedPanel covered 82% of these events. On the other hand, while WES covered all the reported driver SNVs and Indels, SJPedPanel did not cover 7% and 5% of SNVs and Indels, respectively.

**Fig. 4.**
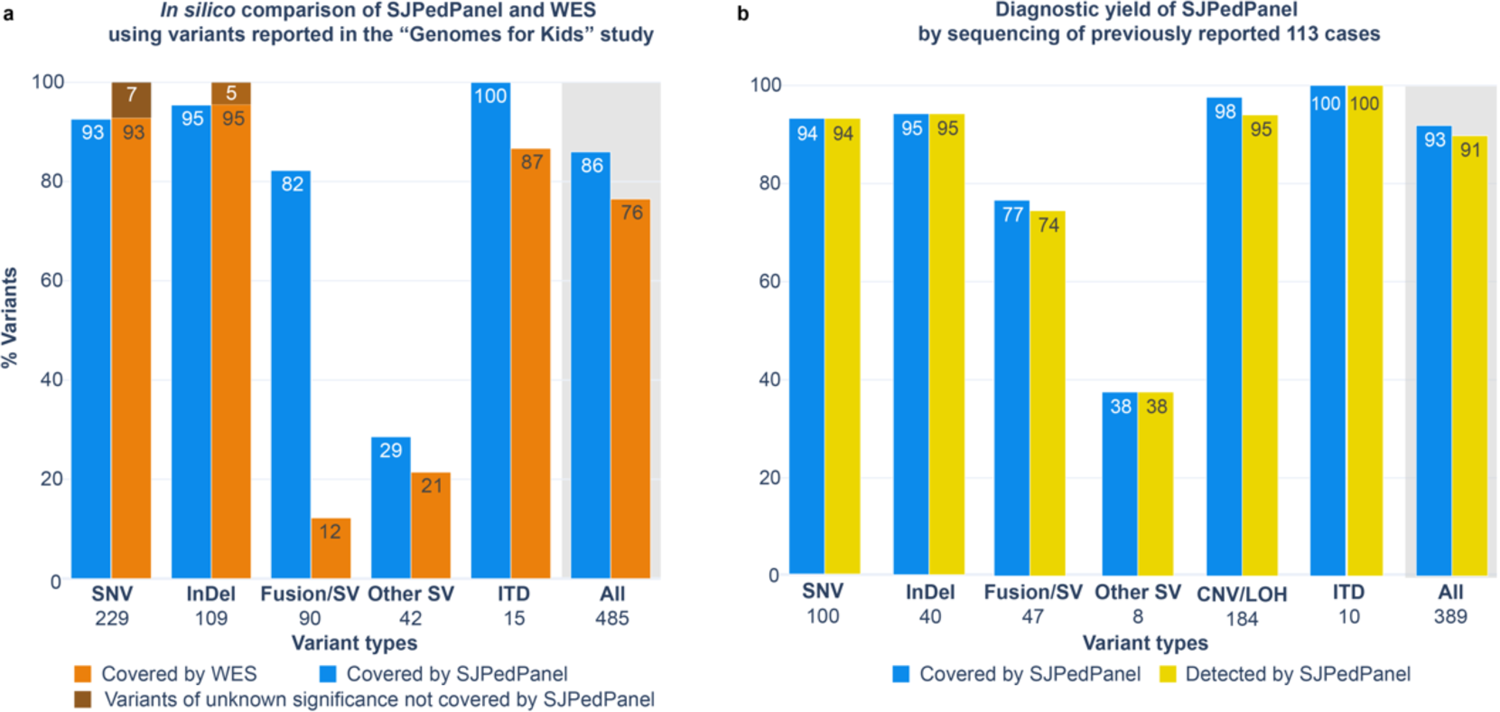
(a) *In silico* coverage comparison between SJPedPanel and WES using percent coverage of variants (SNVs, Indels, Fusion, SV and ITDs) reported in the “Genomes for Kids” study ^71^. The last pair of bars with gray background shows combined percent coverage over “All” 485 variants. **(b) Diagnostic yield of SJPedPanel by sequencing of previously reported 113 cases.** Y-axis shows percentage of covered and detected variants by SJPedPanel over each variant type. The last pair of bars with gray background for “All” variants show combined detection rate. Numbers of reported driver alterations are indicated at the bottom of bars for corresponding variant types.

Interestingly, the host genes of these uncovered SNVs and Indels are rarely mutated (<0.1%) childhood cancers ^2,3^ [**Fig. 4a**, see Additional file 1: Table ST10, “Comment” column]. Of note, SJPedPanel covered 100% of the ITDs, while WES doesn’t cover 13% of these. In fact, the ITDs missed by WES have DNA breakpoints that fall in introns and resulted in duplication of involved exons, such as tandem duplications in *PAX5* ^72^ and *KMT2A* ^73^, for which selected intronic regions were designed in SJPedPanel. Of note, SJPedPanel successfully captured [Additional file 3: Supplementary Fig. S6] the recently described *UBTF* exonic tandem duplications^64^.

We also asked what percentage of patients could benefit from SJPedPanel vs WES. As it turned out, at least one variant per case would have been covered by SJPedPanel in 93% of 210 cases (median: 1, range: 1 to 13), in contrast to 75% of the cases using WES (median: 2, range: 1 to 13) [Additional file 3: Supplementary Fig. S5b]. Most of this gain is due to the capture of intronic breakpoints that result in oncogenic fusions [**Fig. 4a**]. This data demonstrated that SJPedPanel has superior potential for detection and reporting of driver alterations in pediatric tumors than WES.

### Comparison of diagnostic yield between SJPedPanel and WES

To validate above *in silico* findings, we re-sequenced 113 clinical cases with available specimen from previous clinical studies ^4,71^ using SJPedPanel. These samples reflect the broad tumor types and subtypes common in childhood cancer, including 27 hematological malignancies, 43 solid tumors and 43 brain tumors [see Additional file 1: Table ST11 for cohort description]. Common subtypes, such as AML (n=14), ALL (n=10), rhabdomyosarcoma (n=5), neuroblastoma (n=5), osteosarcoma (n=3), Wilms tumor (n=3), high-grade glioma (n=8) and medulloblastoma (n=14) are represented in addition to rare entities, such as melanoma (n=1) and desmoplastic small round cell tumor (n=2) (see Additional file 3: Fig. S7). Among these cases, 389 driver alterations (Methods) are reported via three-platform (WGS, WES, RNAseq) sequencing. These include 100 SNVs, 40 Indels, 55 SVs, 184 CNV/LOHs and 10 ITDs [Additional file 1: Tables ST12-ST16]. Of these, 361 (92.8%) variants were targeted by the panel, including 94 SNVs (94%), 38 Indels (95%), 36 Fusion/SVs (76.59%), 3 other SVs (37.5%), 180

CNV/LOHs (97.82%), and 10 ITDs (100%) [**Fig. 4b**, Additional file 1, Table ST17]. In total 28 P/LP variants were not covered by our panel [Additional file 1, Table ST17]. Of these, 6 were SNVs, 2 were Indels, 11 were Fusion/SVs (not designed), 5 were other SVs and 4 were focal CNVs. The uncovered variants belonged to 26 genes, which are not mutated in published pediatric pan-cancer cohorts ^2,3^ except for *COL1A1* that has a low mutation frequency of 0.2% [see Additional file 1: Table ST4, Additional file 3: Supplementary Fig. S8 and Additional file 1: Table ST17], further supporting their omission from our panel design.

We achieved a mean depth of ∼2500X for all the samples tested [Additional file 1: Table ST18a], which ensures 95% confidence of detection of variants with ≥1% allele fraction (AF) ^74^. As expected, we found an average of 3300 heterozygous SNPs (median: 3405, range: 2227 to 3844) in germline samples (n=30). These samples were pooled as a reference to interrogate CNV/LOH in tumor samples [**Method**; Additional file 1: Table ST18b]. By using a “rotation control” method ^75^ coupled with a recently developed Indel/SV genotyping tool [**Method**], we detected 98% (354 of the 361 covered variants; Additional file 1: Table ST17) of reported driver alterations. SNVs, Indels, ITDs showed detection rate of 100% [**Fig. 4b**]. The Fusion/SVs showed an overall recall rate of 97% (35 out of 36) of the covered alterations. One fusion/SV marker (*RUNX1::RUNX1T1* from the case SJCBF100) that was covered in the panel design with single breakpoint in *RUNX1* was missed due to insufficient depth (21X) of coverage [Additional file 1: Tables ST13 and ST18a], therefore poor capture efficiency in certain genomic regions warrants a future study. By contrast, we could detect 38% (3 out of 8) of other SVs, and the rest of the missed SVs had their breakpoints either in intergenic or non-covered regions, which is consistent with a much larger genomic space for breakpoints in tumor suppressor genes. On the other hand, SJPedPanel detected 94.6% of the reported CNV/LOHs [Additional file 1: Table ST17]. Collectively, SJPedPanel detected 91% of the 389 reported driver alterations from these 113 cases [**Fig. 4b**, pair of last bars with gray background]. Of note, at least one variant was detected for 96.5% cases (n=113), with a median of 3 variants per case (Range = 0-16; Additional file 1: Table ST19). Consistent with the *in-silico* analysis [**Fig. 4a**], a comparison of SNV, Indel, SV and ITD variants (n=205 out of 389) discovered using three platform sequencing^4,71^ (WGS, WES, RNASeq) indicated that SJPedPanel covers 88% of these variants whereas WES covers 78% [Additional file 3: Supplementary Fig. S9]. These data further confirm the superior performance achieved using SJPedPanel for childhood cancers with a panel size approximately 10% the size of WES.

We also highlight the successes and challenges in panel design by using structural variants as examples. First, due to the large genomic size of intronic regions that can be involved in oncogenic fusions, inclusion/exclusion of intronic regions involves a difficult balance between panel size and effective coverage of patient population. In our 2023 study of fusion gene pairs involved in 5,190 childhood cancers ^7^, 72 representative genes are selected for 274 fusion gene pairs and SJPedPanel included 53 of these 72 genes. The maximum mutation frequency of the 19 genes not included [Additional file 1: Table ST3c] in our panel is 0.1% ^7^. Furthermore, inclusion of all relevant introns of all genes involved in oncogenic fusions would need ∼8 Mbp (described in Source data file of Fig, 2k-l mentioned in Liu et al., 2023 ^7^). With the observation that some genes can have multiple fusion partners (e.g., 40 of the 53 included representative genes have between 2 to 32 partner genes), we intentionally left out some partner genes by relying on common representative genes, which reduced the space from ∼8 Mbp (280 introns) to ∼1Mbp (156 introns).

For example, including only the intronic regions of *KMT2A* was sufficient to detect *KMT2A::MLLT1* fusion in pediatric T-ALL patient (SJMLL002) even though we did not include *MLLT1* introns in the panel content [Additional file 3: Fig. S10]. Despite this success, SVs can be challenging to detect. For example, SJPedPanel missed *RUNX1::RUNX1T1* in SJCBF100 because *RUNX1T1* introns were not included (∼100K base pairs needed). As expected, panel also missed an inversion involving *RB1* gene in case SJRB0051 [Additional file 3: Fig. S11] because the DNA breakpoints fall into an intronic region of *RB1* and *RB1* introns are not covered (∼180 kb are needed to cover all *RB1* introns).

Apart from SV/Fusions, SJPedPanel covers well known ITDs such as *FLT3*, *NOTCH1*, *BRAF* etc. [Additional file 1: Table ST16]. SJPedPanel also provides exclusive coverage of *UBTF* gene that was recently described in pediatric AML ^64^. Although in literature *UBTF* ITDs are typically mistakenly detected as small Indels ^3,64,76,77^, our panel successfully detected *UBTF* TDs in two pediatric AML cases (SJAML015373 and SJAML016569; see Additional file 3: Supplementary Fig. S6) ^64^.

### Determining limit of detection

One of the important applications of panel sequencing is disease monitoring, where the tumor burden is typically less than 1% and thus variants are rare and challenging to detect. To investigate the applicability of our panel, we first performed dilution experiments (with 7 tumor concentrations of 10%, 5%, 2.5%, 1%, 0.5%, 0.2%, 0.1%, in addition to pure normal of 0% and pure cancer of 100%) using 6 pediatric cancer cell lines (697, EW-8, K562, ME-1, MOLM-13, and Rh30) and 1 non-cancer cell line (GM12878) as normal control. These 6 lines collectively contain 26 unique P/LP variants (14 SNVs, 4 Indels, and 8 SVs) ^78,79^ [Additional file 1: Table ST20]. The lack of shared driver alterations allowed for pooling of dilutions to reduce the experimental complexity while keeping the diversity of mutation types. For example, at ladder of 0.5%, we mixed 5:5:5:5:5:5:970 cell equivalents from the 6 cancer lines and the normal line, respectively. To ensure sufficient power of detecting variants with low allele fractions, we achieved an average depth of 5,000X for 0.5%, >7,000X for 0.2% and 0.1% dilution concentrations [**Method**, Additional file 1: Table ST21].

We used SequencErr ^80^ and a newly developed SVIndelGenotyper ^81^ (manuscript under review) to perform allele counting followed by variant calling using binomial models with false discovery rate control (**Method**), where the pure normal of 0% was used to estimate background error rates as no cancer-driving somatic alterations are expected in non-cancer cell line GM12878. As seen in **Fig. 5**, the observed allele fractions closely represent corresponding dilution ladders, with R-squared values of 0.7 and 0.72, for biological replicates A and B, respectively [Additional file 1: Table ST22]. Notably, although we achieved >90% detection rate when the dilution concentration is above 0.5%, it diminishes quickly at lower dilutions. At a dilution concentration of 0.1%, recall rate was 69% and 42% in replicates A and B, respectively.

**Fig. 5.**
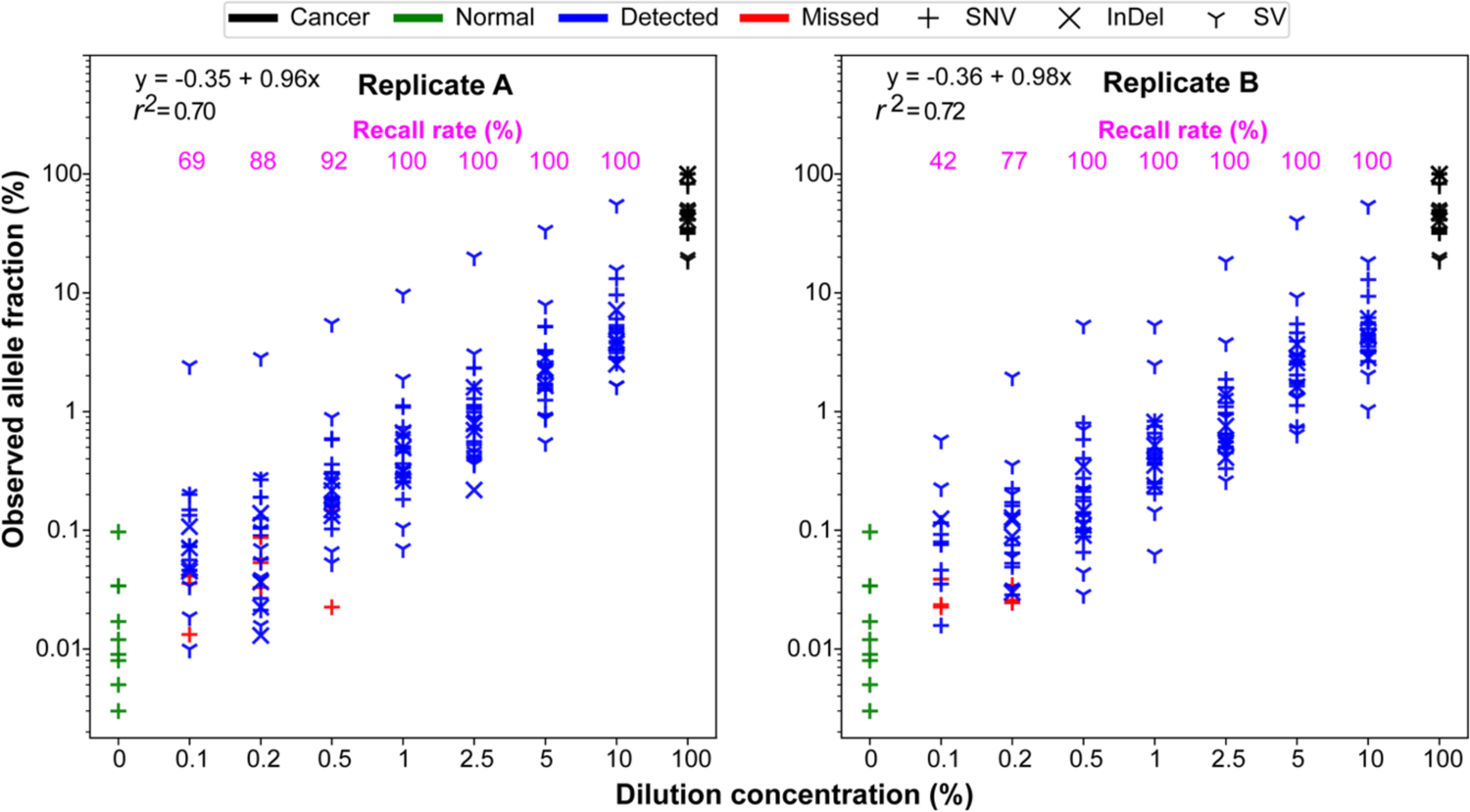
Determining the limit of detection with SJPedPanel. The observed allele fractions of 26 driver alterations are shown on Y-axis as a function of corresponding dilution concentration shown in X-axis. The detection rate for each dilution concentration is shown on top as magenta text. The observed allele fractions of variants from normal and pure tumor cell lines are also shown using green and black points, respectively. The variants detected (*Q* < 0.05) are shown in blue, whereas those missed (*Q* > 0.05) are shown in red. Shown are results from replicate A (a) and replicate B (b).

To further investigate the effect of sequencing depth on detection rate, we performed *in silico* down-sampling experiment [Method; Additional file 1: Table ST23]. For all the dilutions with concentrations ≥ 1%, which were initially sequenced at 5000X and 2500X [Additional file 3: Fig. S12a and S12b], recall rate is found to be close to 100% even after down sampling their depths to 1000X. However, recall rate declined with down sampled depths for dilution concentrations < 1%. For samples with dilution concentration of 0.5% [Additional file 3: Fig. S12b top panel], recall rate dropped from 97% at 3000X to 75% at 1000X. Thus, markers with allele fraction of 0.5% can be reliably detected at 2500X∼3000X, which is concordant with our theoretical binomial calculation of 2,100X [**Method**, **Fig. 5**]. However, for markers with allele fraction of 0.2% and 0.1% the recall rate was <50% even at initial 10,000X data [Additional file 3: Fig. S12c]. This finding suggests that the current LoD is between 0.1% and 0.5% and is consistent with a recent report using cfDNA data ^82^.

The above data suggests a depth of 2,100X will ensures 95% chance of detecting >=5 mutant reads if the true allele fraction is >0.5%, However, considering the sequencing uniformity parameter, where 95% of targeted regions are covered at *μ-* 2*σ* = 0.33*μ*, we would recommend 3 × 2,100 = 6,300X so that >95% of targeted regions will be covered >2,100X and ensure the 95% chance of detecting variants with allele fraction of 0.5%. Since the total space of this panel is ∼0.15% of a human genome, 6,300X corresponds to whole genome sequencing at 9X coverage.

### Case study: Real time tracking of relapsed AML using deep sequencing

To test the suitability of the panel for disease monitoring in a real-world setting, we chose two AML cases (SJAML016582 and SJAML016551) that had remission samples with low tumor purity. These cases have multiple pathogenic and likely pathogenic variants (P/LP) reported by clinical sequencing of diagnosis and relapse samples [Additional file 1: Table ST24] and are ideal for panel sequencing. The diagnosis, remission, and relapse samples were sequenced to an average depth of 5,000X using SJPedPanel per above power calculations.

In case SJAML016582, apart from subtype defining *NUP98*::*NSD1* fusion, 4 pathogenic variants were detected at diagnosis (day 0) and 6 pathogenic variants were detected at relapse (day 315), with 2 variants shared between diagnosis and relapse. With the ultra-deep panel sequencing data, we recovered all variants initially detected by whole genome sequencing, including 4 at diagnosis and 6 at relapse.

Interestingly, panel sequencing detected the SNV that encodes *NRAS* Q61R in diagnosis tumor with allelic fraction (AF) 0.12%, and the MNV that encodes *NRAS* Q61R in relapse tumor with AF 0.07%, both of which are beyond the detection power of whole genome sequencing [Additional file 1: Table ST24-a; **Fig. 6**]. Further, in the day 26 remission data, we detected a high tumor burden (6.56%, Method). Notably, the tumor burden continued to decrease down to AF of 2.88% at day 97 as reflected by the SV responsible for *NUP98::NSD1* fusion.

**Fig. 6.**
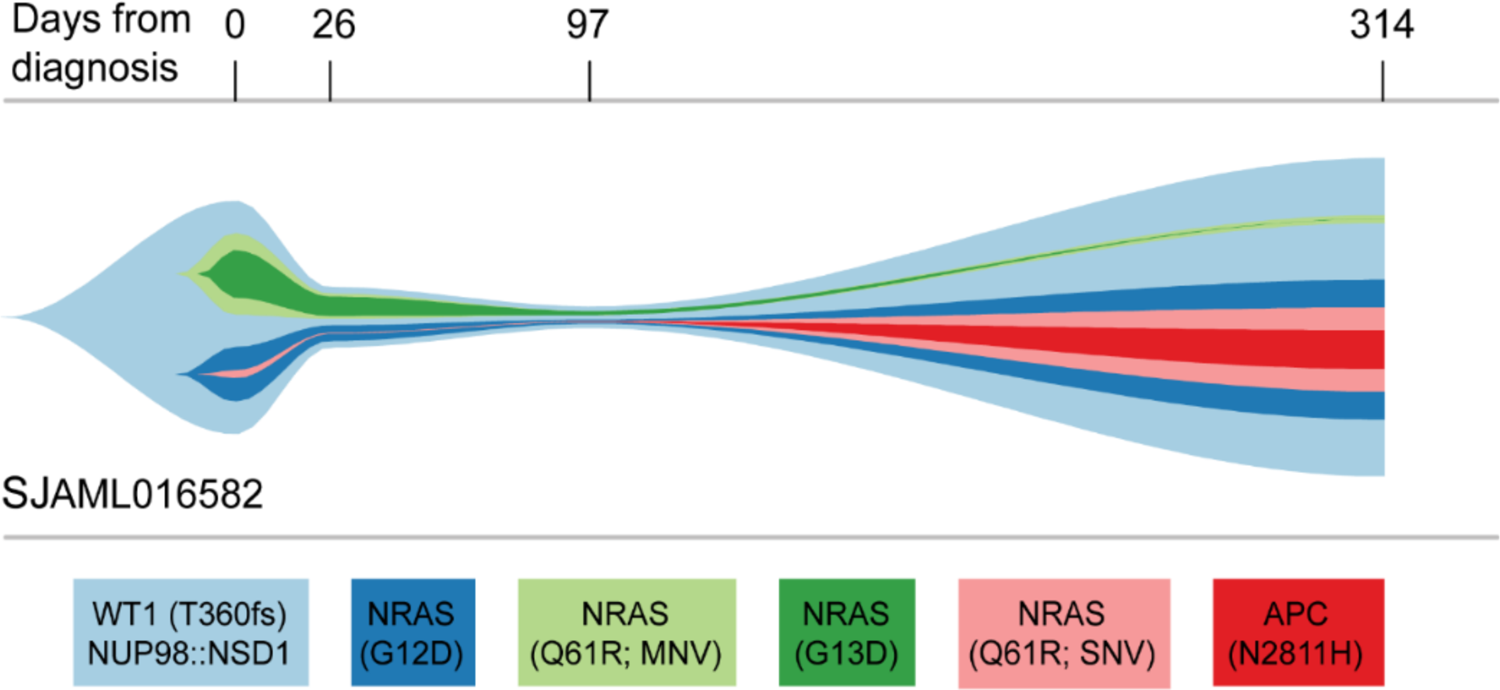
Real time tumor tracking in case SJAML016582. The estimated cellular fractions of subclones at four timepoints from diagnosis (day 0) to relapse (day 314) are shown as a river-plot. Subclones with very low cellularity (e.g., NRAS Q61R (SNV) at diagnosis) were adjusted for visualization purposes using an R package fishplot ^83^. Actual values are available in the Additional file 1: Table ST24-a.

In addition to disease monitoring, we also applied ultra-deep sequencing for detecting measurable residual disease (MRD) to investigate patient response to chemotherapy. Generally flow cytometry is a method of choice for such applications ^84^ in addition to real time PCR ^85^ and droplet digital PCR ^86^. However, these approaches are not as scalable as ultra-deep sequencing. In order to evaluate the efficacy of SJPedPanel to detect MRD, we sequenced three samples from diagnosis (day 0), MRD (day 23) and relapse (day 344) for AML patient SJAML016551. Here the diagnosis and relapse samples are used to define ancestral genetic alterations that are obligated to be present in the MRD sample. For this case, five pathogenic variants, including a *KMT2A::MLLT10* structural variant, were identified to be shared between diagnosis and relapse and are expected to be detected in MRD sample.

Although flow-based MRD detection was negative for this case, our deep sequencing detected all 5 pathogenic variants with allele fraction range between 0.7-1% [Additional file 1: Table ST24-b; Additional file 3: Supplementary Fig. S13]. Together, our data demonstrate the potential ability of SJPedPanel in measuring MRD and monitoring disease progression that could aid in early detection of relapse.

## Discussion

We developed SJPedPanel, a hybridization capture-based assay targeting 357 pediatric relevant genes, including specific oncogenes and tumor suppressors implicated in 44 pediatric cancer genomic studies^2–4,7,26,33,35,61,71^, many through the extensive efforts of the Pediatric Cancer Genome Project ^1^ and NCI TARGET project ^3^, as well as real-time clinical sequencing efforts at St. Jude Children’s Research Hospital employing WGS, WES and RNA-Seq platforms ^4,71^. SJPedPanel includes 297 introns that contain structural variants in 94 genes, accounting for 1.438 Mbp of genomic space, as well as 0.209 Mbp for detecting promoter-hijacking SVs.

Furthermore, 7,590 common SNPs are covered, allowing for the detection of copy number changes and loss of heterozygosity (LOH) at a resolution of ∼1Mb. Due the large differences in the genomic landscape of cancers in children and adults^3^, SJPedPanel has unique coverage of 110 genes frequently implicated in pediatric cancers when compared to other commonly used panels (FoundationOne Heme, FoundationOne CDx, MSK-IMPACT, OncoKids and Oncomine Comprehensive Assay v3). One of the limitations of the current study is that the SJPedPanel was compared with other panels using in silico methods due to limited availability of specimens, which can be addressed in future prospective studies.

We used *in silico* and experimental strategies to evaluate the performance of SJPedPanel for detecting clinically relevant somatic mutations. Using the previously published G4K study, SJPedPanel was found to cover 86% of the reported somatic markers, including SNV, Indel and SV. Similar findings were obtained by using real-time clinical sequencing samples ^4,71^ in which 91% of the reported clinically relevant variants were detected, including all SNVs, Indels and ITDs. At patient level, at least one variant was detected in 96% cases (median 3). These findings establish the ability of SJPedPanel to detect clinically relevant somatic mutations in a wide range of samples from a real-time clinical genomics setting for childhood cancers.

While tumor-normal paired whole genome sequencing remains the gold standard for cancer diagnostics, the overall cost and required bioinformatic pipelines and infrastructure currently limits its broad application. On the other hand, gene-panel based genomics testing can enable many centers to perform NGS-based cancer diagnostics at overall lower cost and faster turn-around time. The content of SJPedPanel allows for more comprehensive detection of the alterations common in pediatric cancer compared to both WES and other panels. An inherent limitation of DNA sequencing panels is the lack of coverage at all critical loci or newly discovered recurrent alterations; however, panel content can readily be updated. For example, the current version of SJPedPanel lacks sufficient coverage to identify the recurrent *ASPSCR1::TFE3* fusion characteristic of alveolar soft part sarcoma or *SSX1*/*SSX2*::*SS18* in synovial sarcoma. Such genes will be incorporated in future versions. To maximize the utility of this panel, the targeted genomic locations are included in Tables ST1 and ST2 [Additional file 1] and future updates will be made readily available to public.

As a proof-of-principle, we demonstrate the application of using SJPedPanel for minimal residual disease (MRD) detection and post-treatment disease monitoring. It would be interesting to see the clinical value of this application using large patient cohorts in the future. Because this is a pan-cancer gene panel for childhood malignancies, it will be relatively straightforward to develop sub panels dedicated to certain cancer subtypes to further reduce the size of the panel, and in turn enabling much higher depth with a similar cost. We expect SJPedPanel to significantly impact the management and diagnosis of children with cancer.

## Material and Methods

### Panel design

Based on extensive research and literature review of pan-cancer genome profiling studies a list of exonic and/or intronic regions (n=5,009 regions, 2.82 Mbp) from 357 genes that are frequently implicated in pediatric cancers was compiled to detect single nucleotide variants (SNVs), small insertions & deletions (Indels), gene fusions, structural variants (SVs), and internal tandem duplications (ITDs). We also curated a list of 7,590 single nucleotide polymorphic markers (SNPs) evenly spread across human chromosomes to detect large genomic structural rearrangements such as copy number variations (CNVs) and loss of heterozygosity (LOH). The details of all the genomics regions and SNPs used to assemble the pediatric pan cancer, termed as SJPedPanel, are available in Additional file 1: Tables ST1 and ST2.

### Capture efficiency of the SJPedPanel

We generated one high-depth (∼2000X) and one low-depth (∼200X) targeted sequencing libraries (two replicates) using SJPedPanel using COLO829BL cell line (a gold-standard cell line for clinical validation and other established studies ^69,70^). These libraries were sequenced on Illumina NovaSeq and NextSeq platforms, respectively. The data generated was used to evaluate the capture performance of the probes and uniformity of coverage across regions and loci of the SJPedPanel [**Fig. 3** and Additional file 3: Fig. S2].

### Dilution experiment

A dilution experiment using 6 cancer cell lines (ME-1, 697, Rh30, EW-8, K562, Molm13, courtesy of Elizabeth Stewart, Department of Oncology, St. Jude Children’s Research Hospital) and a non-cancer cell line (GM12878) was designed to achieve seven tumor concentrations with two replicates each. The seven dilutions were divided in three groups a) ultra-low (0.1%, 0.2%), b) low (0.5%, 1%), and c) medium (2.5%, 5% and 10%), which were sequenced at depths of 10,000X, 5000X and 2500X respectively. The cell lines were also sequenced independently in undiluted forms at 25,000X to estimate the original allele fractions of 26 cell-line-specific markers (14 SNVs, 4 Indels, 8 SVs; all these markers are confirmed to be exclusively detected from one of the six cell lines) given in Additional file 1: Table ST20. Recall rate of these known markers across different dilutions was used to assess the limit of detection of the SJPedPanel.

The limit of detection (LoD) is determined by two critical factors: 1) the sequencing depth (also known as power) and 2) the noise level. For example, if the true allele fraction is 1%, a sequencing depth of 913X will ensure 95% chance of detecting this variant with >=5 mutant alleles ^74^. In consideration of the high range of dilution concentrations, we aimed for 2,500X depth for dilution ladders >1%, 5,000X for ladders 0.5% and 1%, and 10,000X for ladders 0.1% and 0.2%. On the other hand, the noise level is typically regarded as background error rate. Mutations with higher background error rates are more difficult to detect because the true signal can be overwhelmed by the background noises. We previously developed computational error suppression methods to achieve error rate of ∼10^-6^-10^-4^ for substitutions ^87^ and similar methods and results have been achieved for Indels and SVs (manuscript under review).

### *In silico* down sampling experiment

We performed *in silico* down sampling of data from a set of cancer cell line dilution samples to find out the trade-off between recall rate, down sampled depth of sequencing and associated cost estimates. The samples originally sequenced at 2,500X were further down sampled to simulate depths of sequencing at 1,000X, 1,500X and 2,000X, whereas the samples which were sequenced at 5,000X and 10,000X were down sampled to simulate depths of sequencing at 1,000X, 1,500X, 2,000X, 3,000X. For each of the desired down sampled depth, 10 samples were simulated, each consisting of randomly sampled reads at loci of 14 SNVs. These simulated samples were used to determine the trade-off between recall rate and depth of sequencing.

### Investigating diagnostic yield using clinical sample re-sequencing

Based on sample availability we selected 113 specimens from previously sequenced pediatric cancer cases treated at St. Jude Children’s Research Hospital to represent a wide range of cancer subtypes common in pediatrics. Samples were chosen primarily from the pilot study cohort (n=40) ^4^ and G4K studies (n=73) ^71^ and had previously reported clinically relevant markers identified by triple platform approach of whole genome, whole exome, and transcriptome sequencing. The recall rate of clinically relevant markers from these cases was used to establish the diagnostic yield of the SJPedPanel. Here seven markers from 3 hypermutator cases (SJHGG030335, SJHGG030336 and SJST030211) were downgraded to VUS (variant of unknown significance) for this analysis per communication with corresponding author of the G4K study [see “Comment” column in the Additional file 1: Table ST12], resulting in 140 of SNV/Indel, 55 of fusion/SVs, 184 CNV/LOH and 10 ITDs (total = 389). A list of cases and their cancer subtypes used for these purposes is provided in the [Additional file 1: Table ST11].

### Library preparation, capture, and sequencing

DNA samples were obtained and subjected to DNA-seq library preparation and target enrichment followed by sequencing in the Clinical genomics laboratory as described below. An input of 100ng of DNA was used to construct libraries using the Twist Library Preparation Enzymatic Fragmentation (EF) Kit 2.0 (Twist Biosciences, CA) following the manufacturer’s instructions. Capture oligos were designed to detect putative SNVs, Indels, SVs, ITDs and CNVs in 357 genes of clinical interest. SJPedPanel was synthesized at Twist Biosciences and is described in detail in the section on panel design. Eight libraries were pooled at a time and target enrichment for the SJPedPanel baits was carried out using Twist hybrid capture protocol following manufacturer’s instruction. Paired-end 150-cycle sequencing was performed on NovaSeq or NextSeq instruments (Illumina Inc, CA) as appropriate. Where necessary, additional sequencing (“top off”) was performed to ensure that a sequencing depth of at least 1000X was achieved in all cases.

### Early detection of relapsed AML cases

To test the panel’s capability for disease monitoring, two pediatric AML cases (SJAML016582 and SJAML016551) with material available at diagnosis, relapse, and remission timepoints were analyzed. Both samples provide multiple trackable somatic markers, including structural variants and SNVs. Samples were subjected to deeper sequencing depths of >5000X after targeted capture to ensure detection of low-level variants at <1%. Average of variant allele fractions (VAF) of detected somatic variants were used to estimate tumor burden at corresponding time points.

### Coverage comparison between SJPedPanel and Whole exome sequencing

The content of SJPedPanel was compared with that of whole exome sequencing (WES) manifest to highlight the differences in coverage of hg19 genomic regions. An Illumina Exome 2.0 Plus hg19 BED file ^88^ padded with 10 bp was used for region intersection analyses. We utilized recently reported somatic variants from the Genomes for Kids (G4K) ^71^, a real-time three-platform sequencing study of 309 pediatric cancer patients to benchmark the coverage of reported pathogenic and likely pathogenic variants between SJPedPanel and WES.

### Comparison of SJPedPanel with other panels

We compared the content of the SJPedPanel with DNA content of five other commercially available panels including FoundationOne Heme ^57^, FoundationOne CDx ^58^ by Foundation Medicine Inc., MSK-IMPACT ^6,59^ by Memorial Sloan Kettering Cancer Center, OncoKids ^5^ by Children’s Hospital Los Angeles and Oncomine Comprehensive Assay v3 by Thermo Fisher Scientific Inc. ^60^. These panels collectively represent the breadth and diversity of clinical gene panels. Since most of the providers do not provide the exact coordinates of the regions in the panel, we compared the content of these panels using standardized gene names with the help of official gene symbols and synonyms from the NCBI gene database ^89^.

### Bioinformatics analyses

The adapter trimmed paired end FASTQ files generated on the Illumina NovaSeq/NextSeq platforms were assessed for sequence and instrument quality using FastQC v0.11.9 ^90^ and SequencErr v2.0.9 ^80^. The reads were mapped against GRCh37 build using BWA aln 0.7.12-r1039 ^91^. The utility commands in SAMtools v1.7 ^92^ and BEDTools v2.25.0 ^93^ were used perform simple operations using BAM and BED files. The count files obtained from BAM files using SequencErr were further passed as an input to DeepSeqCoverageQC v0.3.1 ^94^ to compute depth of coverage QC metrics of the sequenced samples over loci/regions of the SJPedPanel. The genotyping of SVs and Indels to compute the allele fractions was carried out using SVIndelGenotyper ^81^. The CNVs were detected using CNVkit v0.9.10 ^95^. The BAM files of 30 germline samples were used to create a pooled reference of per-bin copy number estimates. The segment & bin-level call files along with CNV diagrams generated by CNVkit batch command were used to review the CNV calls in tumor samples. To determine the LOH in sequenced samples, the minor allele frequencies of 7590 SNPs were computed using count files generated by SequencErr and subsequently used to generate allelic imbalance plots over chromosomes. The output files and diagrams generated by CNVkit v0.9.10 and the allelic imbalance figures used to review CNV and LOH events are available from Zenodo repository ^96^.

A previously developed “rotation control” method ^75^ was used to obtain the background count of the variants for binomial testing and Q-values ^97^ were used to assess the statistical significance of detection based on binomial testing. Briefly, frequency of any variant from the original mutation-positive sample was considered as foreground (*V_f_*) and frequency of the same variant derived from the aggregated counts from rest of the samples, which were expected to be wild type in the original analysis, was considered as background or control (*V_b_*) for the binomial testing. Thus, the background counts were rotated w.r.t. change in the foreground variant and sample of interest. A sample is called mutation-positive if the Q-value was found to be < 0.05. All the statistical analyses were performed using R v4.0.3 ^98^.

## Data Availability

All data produced in the present study are available upon reasonable request to the authors. The cell line data produced are available online at the European Nucleotide Archive (ENA) at EMBL-EBI under accession number PRJEB64356 (https://www.ebi.ac.uk/ena/browser/view/PRJEB64356).

https://github.com/pandurang-kolekar/DeepSeqCoverageQC

https://github.com/stjude/SVIndelGenotyper

https://zenodo.org/doi/10.5281/zenodo.8173838

https://www.ebi.ac.uk/ena/browser/view/PRJEB64356

## Declarations

### Availability of data and materials

The cell line data generated for this study have been deposited in the European Nucleotide Archive (ENA) at EMBL-EBI under accession number PRJEB64356 (https://www.ebi.ac.uk/ena/browser/view/PRJEB64356). The accession numbers of the samples are listed in Additional File 1: Tables ST5 and ST21.

### Authors’ contributions

X.M. and J.M.K. conceived the research. P.K. collected data, performed data analyses, generated figures and implemented the *DeepSeqCoverageQC* algorithm. V.B. and L.D. performed sequencing experiments. J.M.K. coordinated clinical specimen acquisition. Y.L. and Q.T. implemented *SVInDelGenotyper* algorithm. P.K., V.B., L.D.,Y.L.,S.F.,Q.T.,H.M.,A.L.H,E.P.,Z.L.,J.M.,J.N,J.G,J.M.,D.P.,S.M.,L.W., E.S.,J.E., J.M.K. and X.M. contributed to analyses. All authors read and approved the final manuscript.

### Authors’ information

Pandurang Kolekar, Vidya Balagopal and Li Dong contributed equally to this work.

### Ethics approval and consent to participate

This study was approved by the St. Jude institutional review board (IRB) and informed consent was obtained for samples collection from the patient, parents or guardians. Subjects were not compensated for participation. All patient samples are de-identified.

## Acknowledgements

X.M and J.M.K thank Elizabeth Stewart for providing cell lines for our dilution experiments.

