## Additional file 2 for "SJPedPanel: A pan-cancer gene panel for childhood malignancies"

Chromosomal distribution of genes (n=357) in SJPedPanel

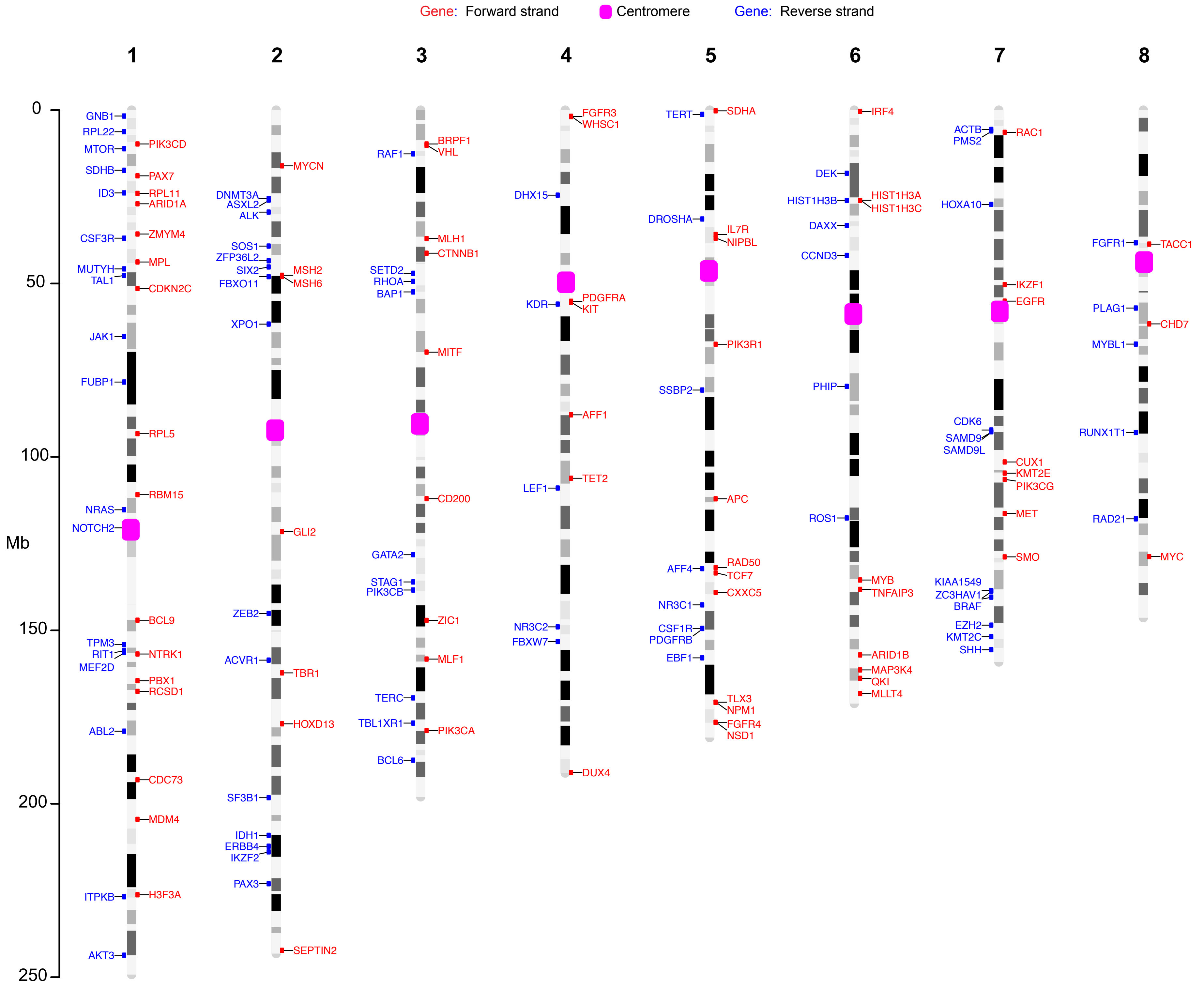

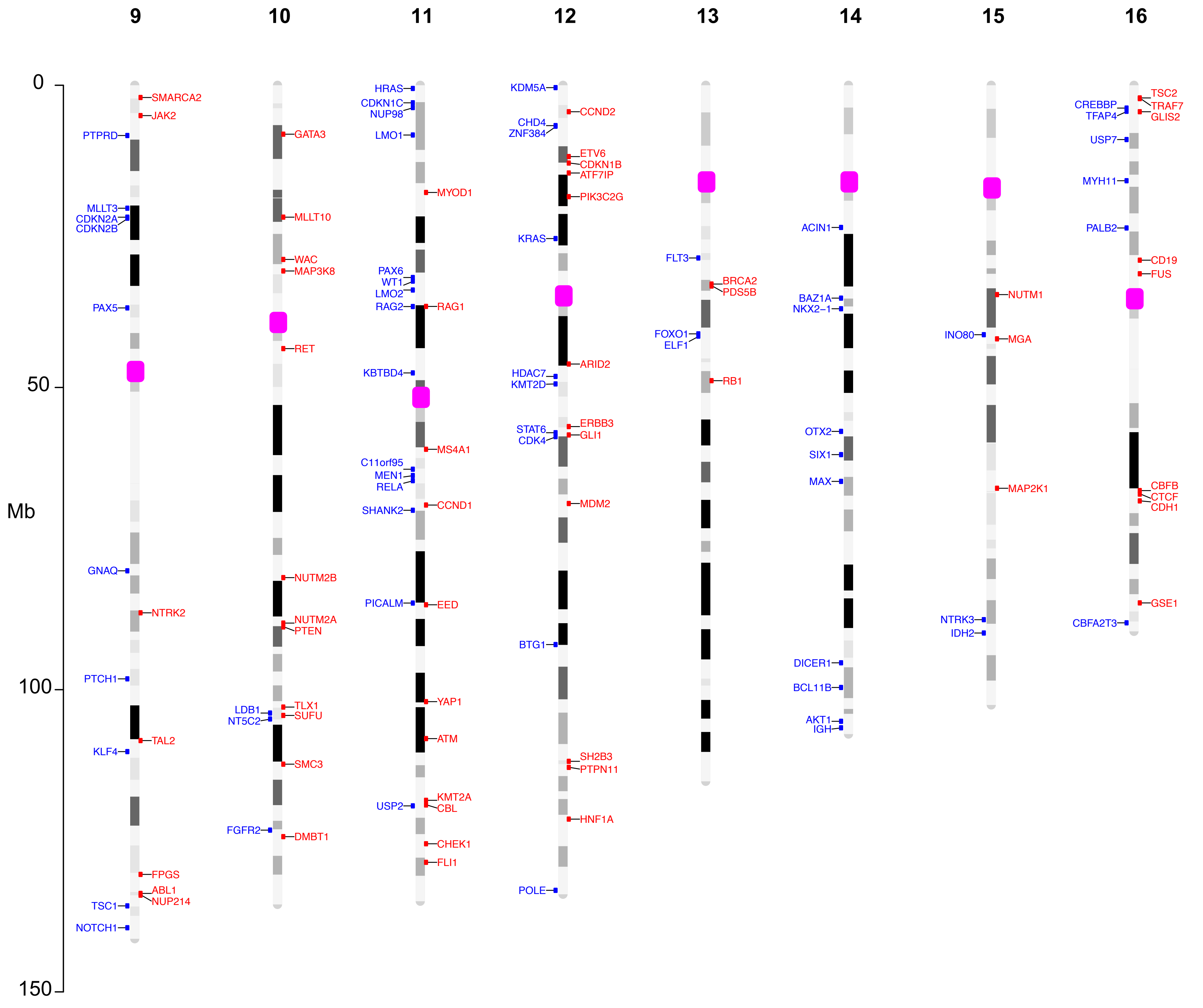

9
10
11
12
13
14
15
16
0
50
100
150
Mb
SMARCA2
JAK2
PTPRD
MLLT3
CDKN2A
CDKN2B
PAX5
GNAQ
NTRK2
PTCH1
TAL2
KLF4
FPGS
ABL1
NUP214
TSC1
NOTCH1
GATA3
MLLT10
WAC
MAP3K8
RET
NUTM2B
NUTM2A
PTEN
TLX1
SUFU
LDB1
NT5C2
SMC3
FGFR2
DMBT1
HRAS
CDKN1C
NUP98
LMO1
MYOD1
PAX6
WT1
LMO2
RAG2
RAG1
KBTBD4
MS4A1
C11orf95
MEN1
RELA
CCND1
SHANK2
PICALM
EED
YAP1
ATM
USP2
KMT2A
CBL
CHEK1
FLI1
KDM5A
CCND2
CHD4
ZNF384
ETV6
CDKN1B
ATF7IP
PIK3C2G
KRAS
ARID2
HDAC7
KMT2D
STAT6
CDK4
ERBB3
GLI1
MDM2
BTG1
SH2B3
PTPN11
HNF1A
POLE
FLT3
BRCA2
PDS5B
FOXO1
ELF1
RB1
ACIN1
BAZ1A
NKX2−1
OTX2
SIX1
MAX
DICER1
BCL11B
AKT1
IGH
NUTM1
INO80
MGA
MAP2K1
NTRK3
IDH2
TSC2
CREBBP
TRAF7
TFAP4
GLIS2
USP7
MYH11
PALB2
CD19
FUS
CBFB
CTCF
CDH1
GSE1
CBFA2T3

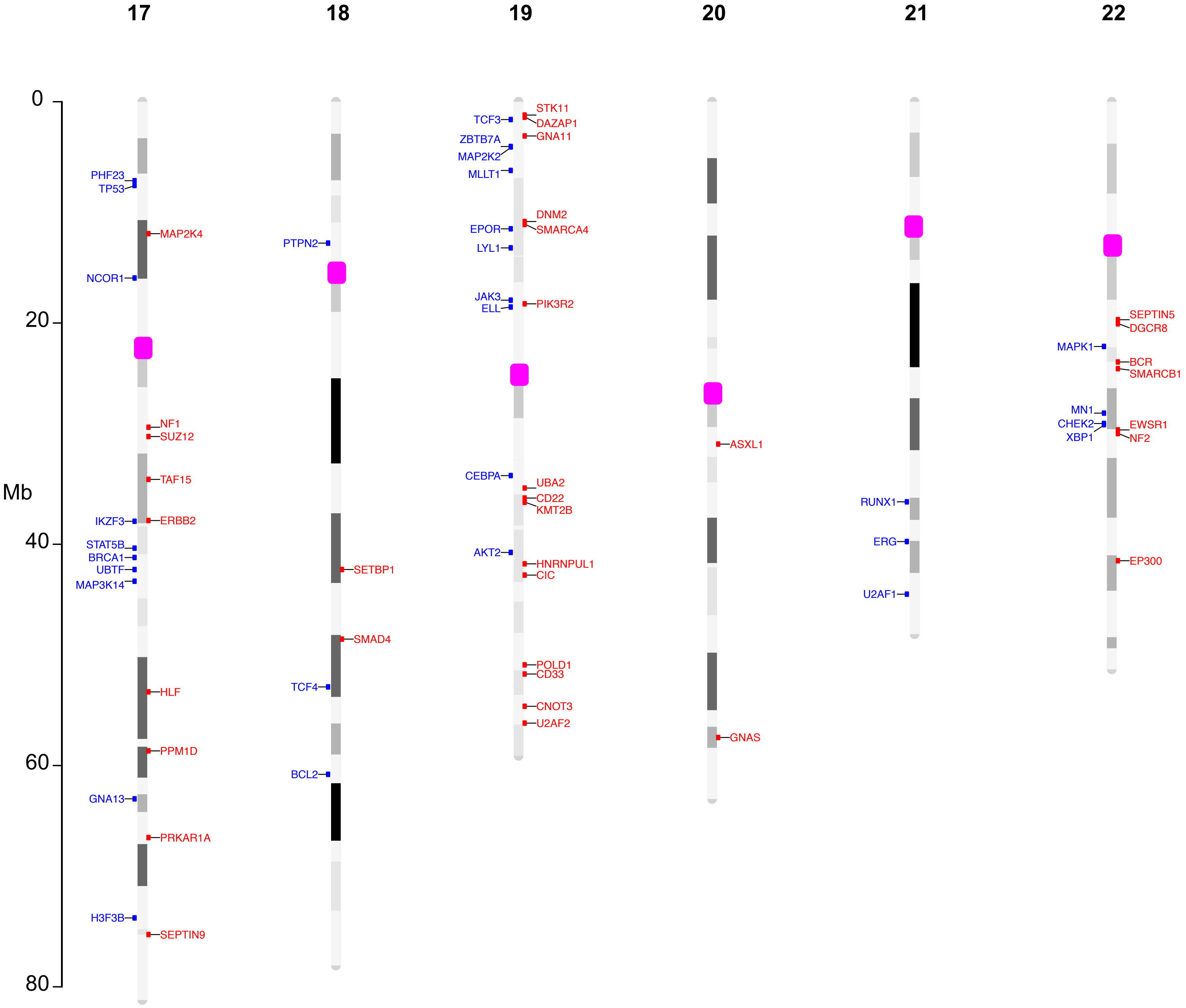
17
18
19
20
21
22
0
20
40
60
80
Mb
PHF23
TP53
MAP2K4
NCOR1
NF1
SUZ12
TAF15
IKZF3
ERBB2
STAT5B
BRCA1
UBTF
MAP3K14
HLF
PPM1D
GNA13
PRKAR1A
H3F3B
SEPTIN9
PTPN2
SETBP1
SMAD4
TCF4
BCL2
TCF3
STK11
DAZAP1
GNA11
ZBTB7A
MAP2K2
MLLT1
DNM2
SMARCA4
EPOR
LYL1
JAK3
ELL
PIK3R2
CEBPA
UBA2
CD22
KMT2B
AKT2
HNRNPUL1
CIC
POLD1
CD33
CNOT3
U2AF2
ASXL1
GNAS
RUNX1
ERG
U2AF1
SEPTIN5
DGCR8
MAPK1
BCR
SMARCB1
MN1
CHEK2
XBP1
EWSR1
NF2
EP300

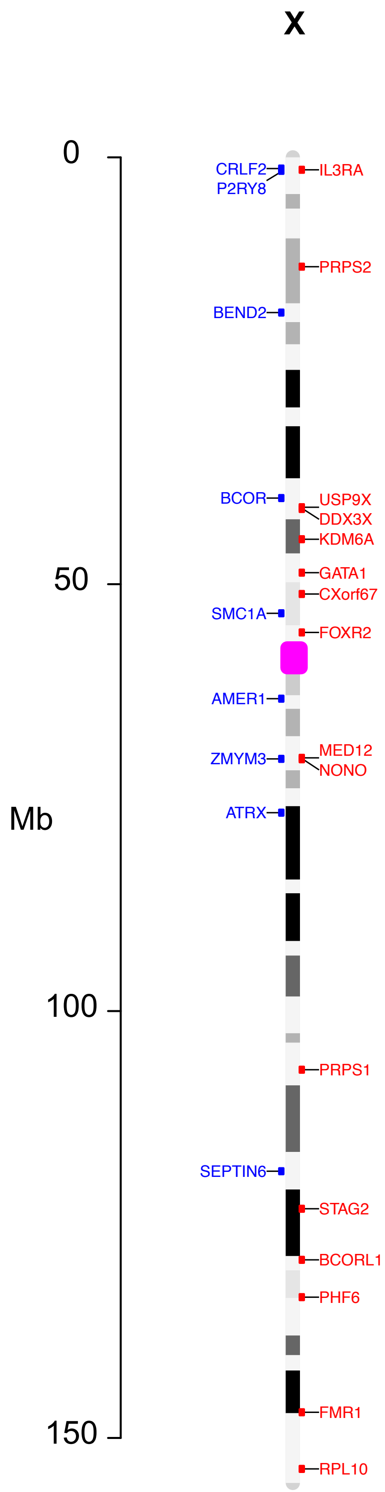
X
0
50
100
150
Mb
CRLF2
P2RY8
IL3RA
PRPS2
BEND2
BCOR
USP9X
DDX3X
KDM6A
GATA1
CXorf67
SMC1A
FOXR2
AMER1
MED12
ZMYM3
NONO
ATRX
PRPS1
SEPTIN6
STAG2
BCORL1
PHF6
FMR1
RPL10
