## Additional file 3: Supplementary figures for "SJPedPanel: A pan-cancer gene panel for childhood malignancies"

*: Contributed equally

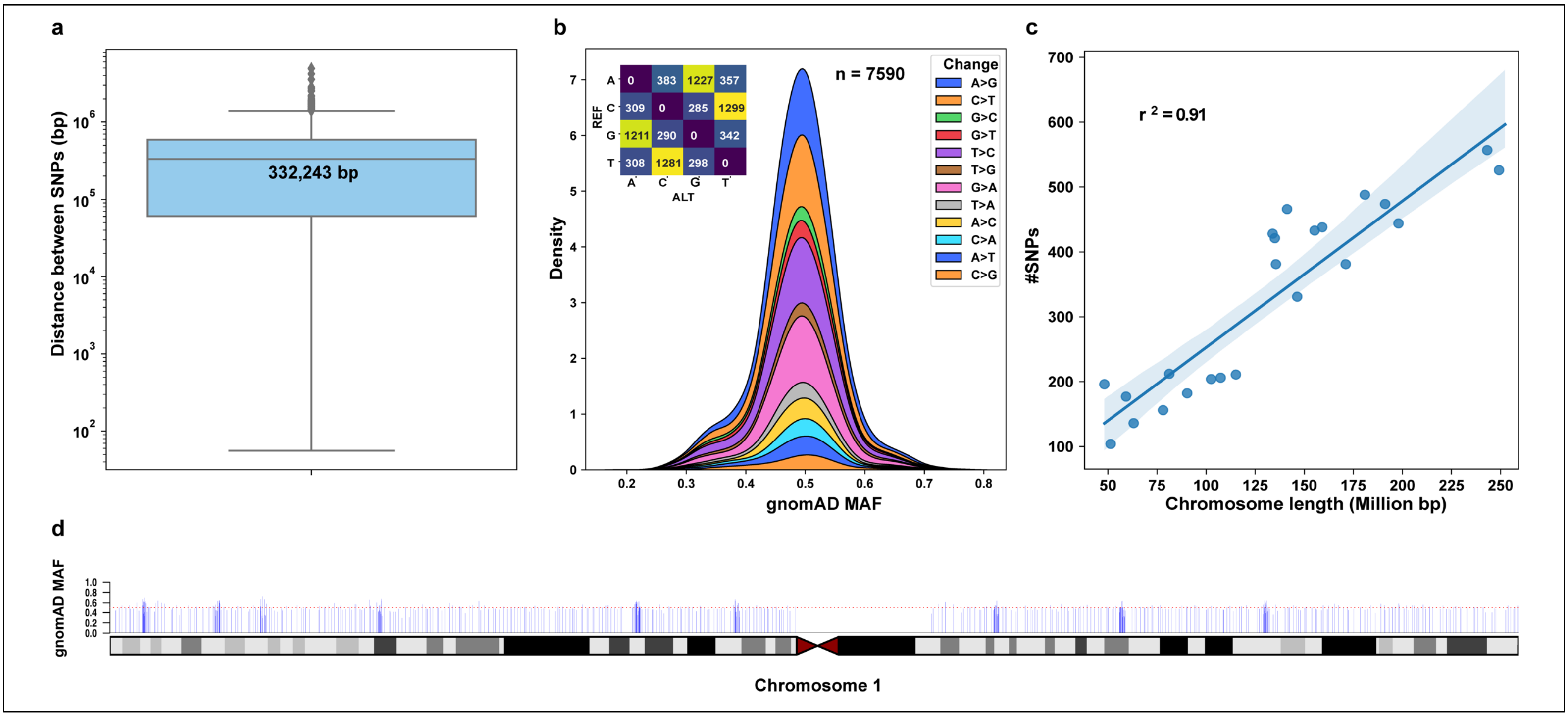


**Figure S1 – Summary of 7590 SNPs included in the SJPedPanel**

(a) Boxplot of distance (base pairs, bp) between SNPs.
(b) Distribution of minor allele frequencies of SNPs as per gnomAD v2.1.1 database available from https://gnomad.broadinstitute.org/
(c) Correlation between length of chromosomes (n=23) and number of SNPs on respective chromosomes
(d) Representative plot showing evenly spread locations of SNPs (n=528) over the length of chromosome 1 (249.25 Mbp; hg19). The Y-axis shows minor allele frequencies of SNPs as per gnomAD v2.1.1. Plots for all the chromosomes available in the Additional file 4.

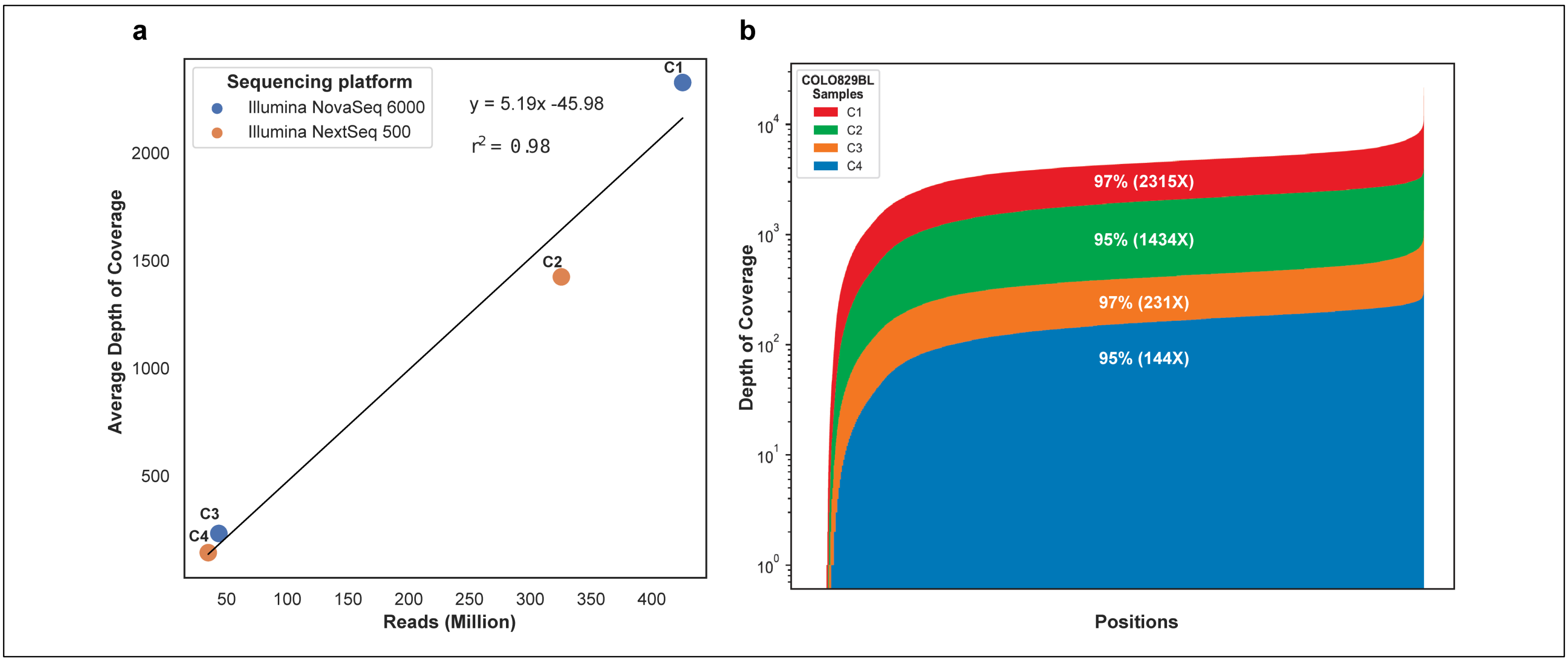


**Figure S2 – Target Capture performance of SJPedPanel using COLO829BL cell line samples (C1-C4)**

(a) Correlation between average depth of coverage and number of reads.
(b) Uniformity of coverage across panel position (2.82 Mbp). The % values indicate uniformity of coverage whereas numbers in the parentheses indicate average depth of coverage of the sample excluding observations in upper and lower 2.5 percentiles.

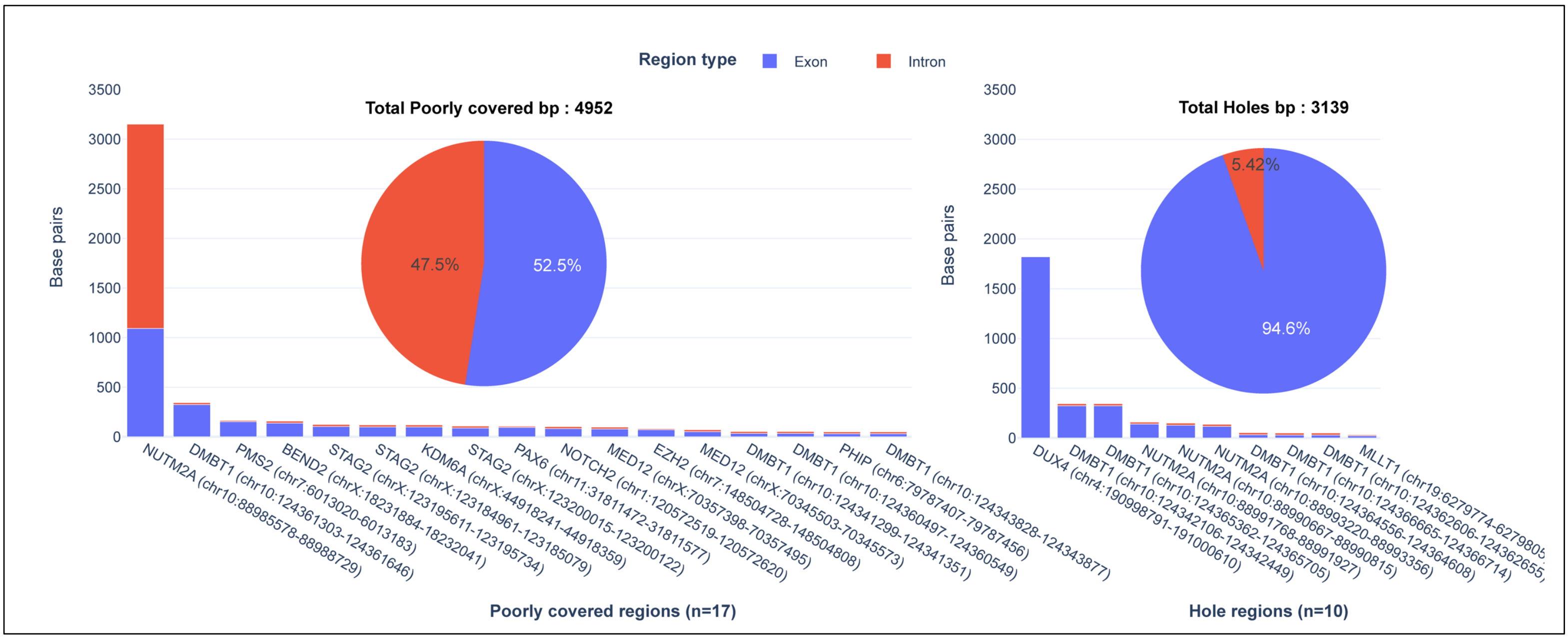


**Figure S3 – Composition of 27 consistently poorly covered and hole regions in the SJPedPanel.**

The X-axis shows names and coordinates of 27 regions split between poorly covered (n=17, left) and hole regions (n=10, right). Pie chart shows overall distribution of bases from exon/intron parts of the regions. Stacked bars show the same distribution of bases over respective regions. Total bases occupied by each group of regions is shown above the Pie charts (4952 + 3139 = 8091 bp).

**Note:** The small intronic portions shown above the bars is contributed by the padded portions on either ends of exons.

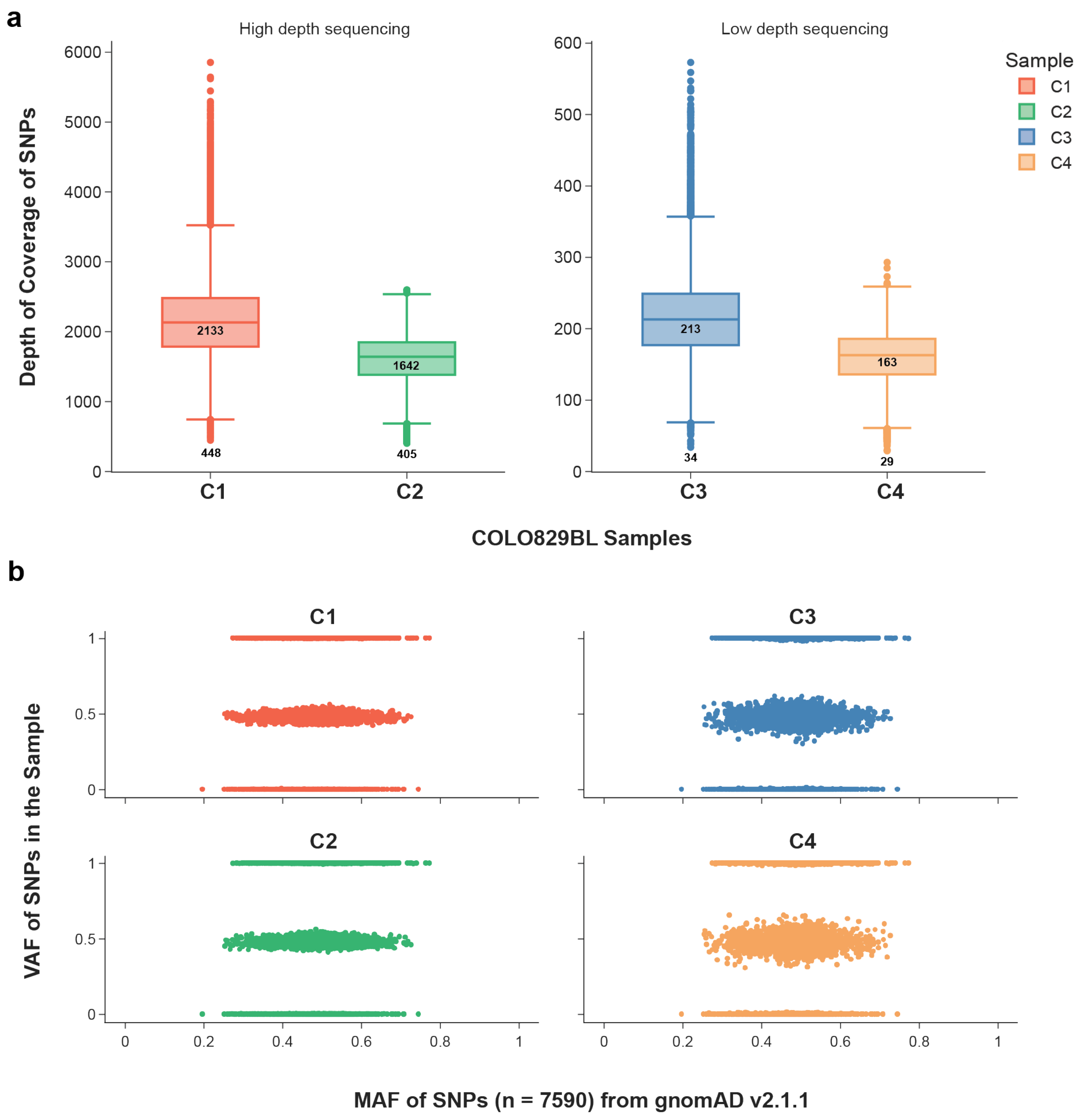


**Figure S4 – Capture performance of 7590 SNPs in SJPedPanel using COLO829BL cell line samples (C1-C4)**

**(a)** Depth of coverage of SNPs (n=7590) in high depth sequencing (C1, C2) and low depth sequencing (C3, C4) samples. Minimum and median depth of coverage values are shown along the axis of boxplots. **(b)** Variant allele frequencies (VAF) of SNPs (n=7590) in COLO829BL samples (C1-C4) are shown on Y-axis and MAFs for those SNPs in gnomAD v2.1.1 (https://gnomad.broadinstitute.org/) are shown on X-axis of scatter plots. **Note:** Depth of coverage and VAF values for these samples are provided in Additional file 1: Supplementary table ST9.

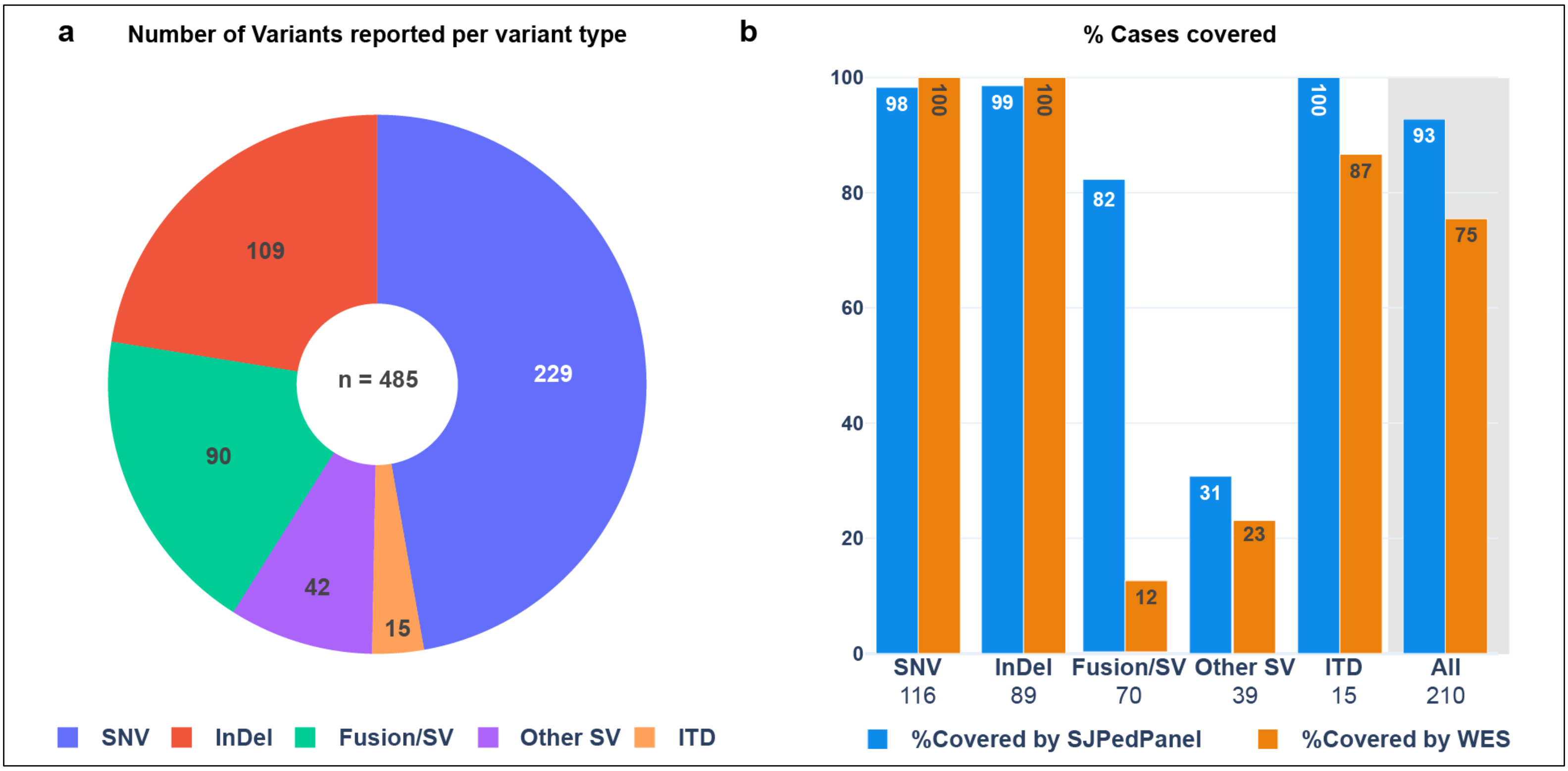


**Figure S5 – *In silico* comparison of SJPedPanel and WES using variants reported in “Genomes for Kids” study.**

(a) Number of variants per variant type (SNV, InDel, Fusion/SV, Other SV and ITD) reported in the G4K study (n=485)

(b) Percentage of cases covered per variant type by SJPedPanel and WES considering coverage of at least one variant per case. Number of total cases represented by the variant type are given below respective bars. The last pair of bars with gray background for “All” variants collectively show % of 210 unique cases covered represented by all the variants (n=485)

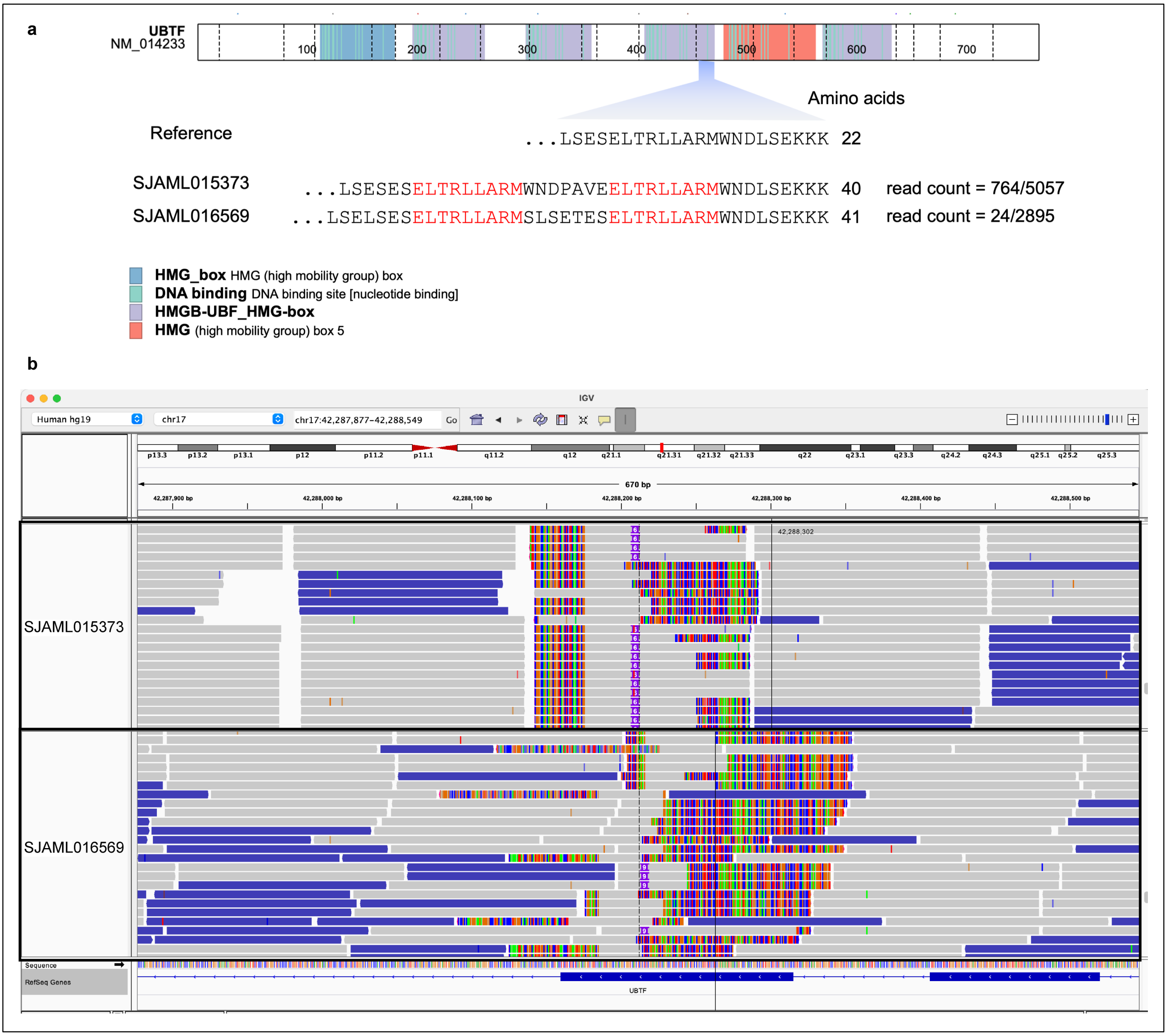


**Figure S6 – UBTF-TDs in pediatric AML cases (SJAML015373 and SJAML016569) detected using SJPedPanel.**

**(a)** Figure shows an illustrative schema of *UBTF* protein and amino acid sequences within the HMG domain 4 of both UBTF-wild-type and UBTF-TDs. A part of UBTF-TDs encoded on exon 13 of *UBTF* gene is shown in comparison with UBTF-wild-type of human. Amino acid sequences highlighted in red denote leucine-rich sequences duplicated in UBTF-TDs for both the cases. **(b)** Sequence reads from the samples SJAML015373 (top track) and SJAML016569 (bottom track) aligned against the human genome (hg19) consisting of UBTF-TDs are shown IGV genome browser (https://igv.org/)

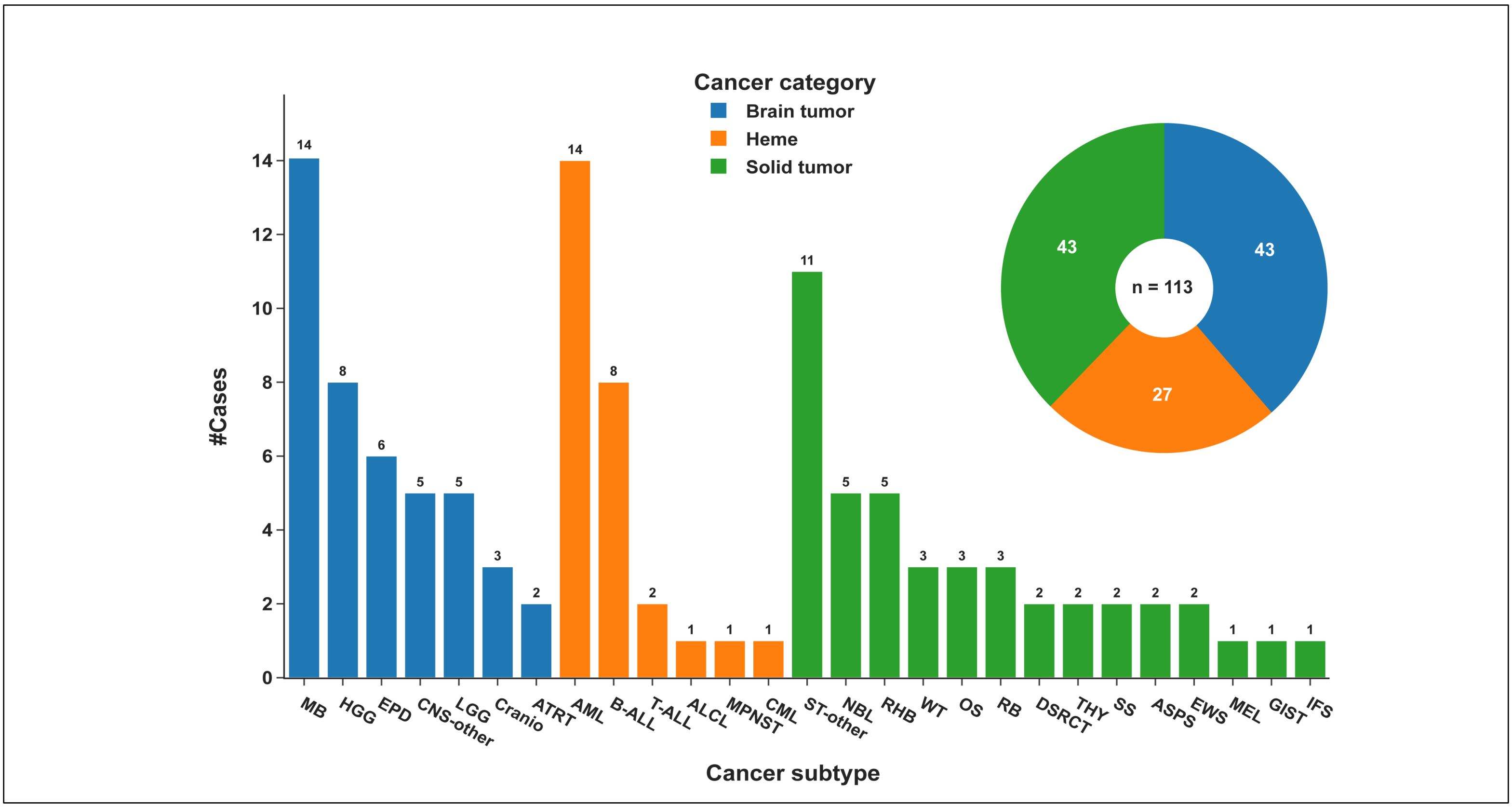


**Figure S7 – Spectrum of cancer subtypes under different cancer categories represented by cases (n=113) chosen to determine diagnostic yield of the panel.**

A donut pie chart shows number of cases under 3 major cancer categories (Brain tumor, Heme and Solid tumors). The bar chart shows number of cases for each cancer subtype within respective cancer categories. The description of cases and cancer subtypes is available in the Additional File 1: ST11.

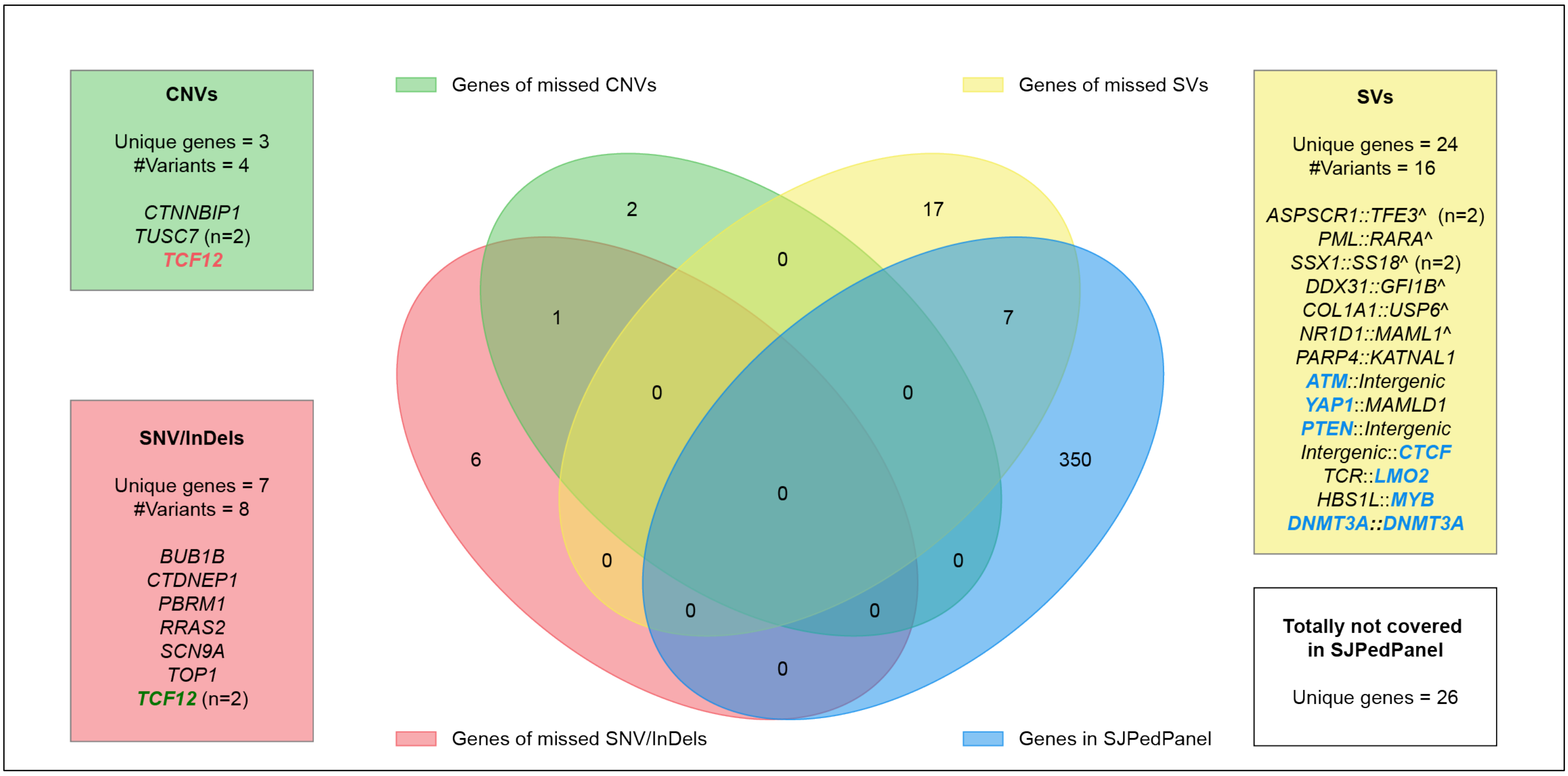


**Figure S8 – Summary of 28 variants not targeted by SJPedPanel and missed as part of diagnostic yield.**

Venn diagram shows intersection of genes of 28 non-covered variants (SNV/InDels: 8, SVs: 16 and CNVs: 4). Names of the unique genes are shown in accompanying boxes color coded as per the variant group. Genes of seven of SV partners were covered in the SJPedPanel except for the concerned SV breakpoint(s) hence missed the respective SVs. Uniquely 26 genes were totally not covered by the SJPedPanel.

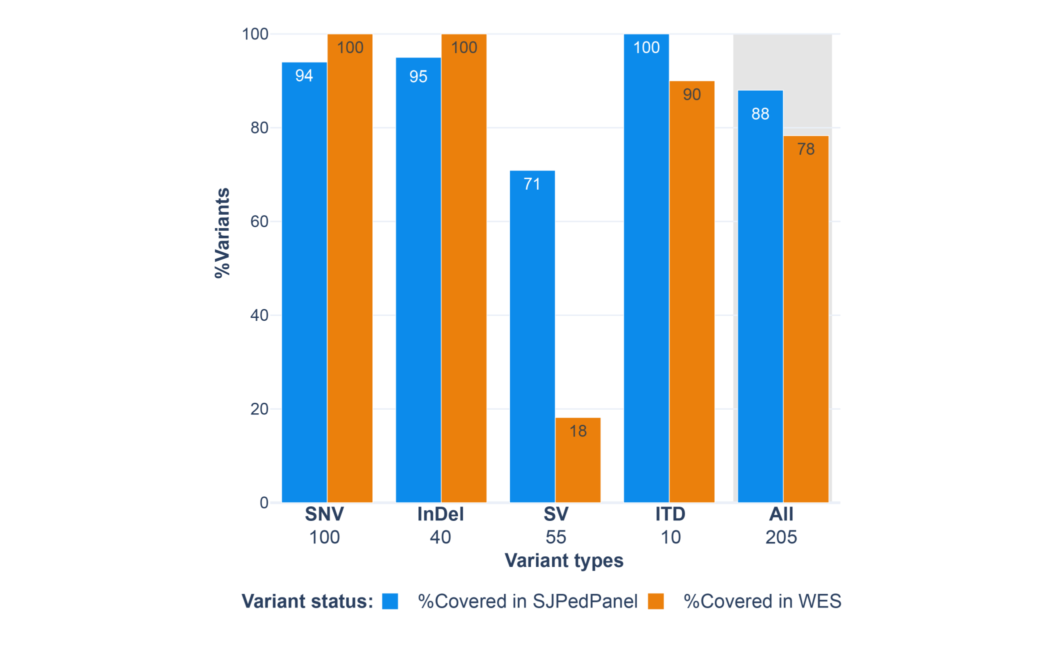


**Figure S9 – Comparison of coverage between SJPedPanel and WES using subset of 205 variants as part of diagnostic yield.**

Percentage of variants covered per variant type by SJPedPanel and WES considering SNV, InDel, SV and ITD events. Number of total variants by the variant type are given below respective bars. The last pair of bars with gray background for “All” combined variants show % of 205 variants covered by SJPedPanel and WES.

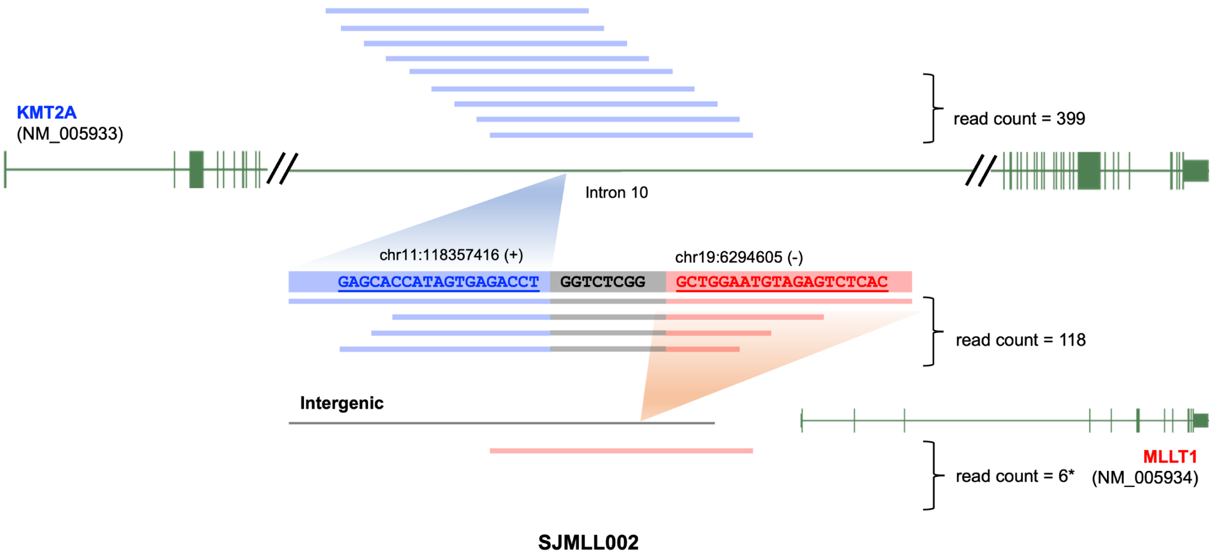


**Figure S10 – Detection of complex structural arrangement using SJPedPanel *KMT2A::MLLT1* fusion in pediatric T-ALL patient (SJMLL002)**

Gene structure for *KMT2A* (NM_005933) is shown on top. A portion of intron 10 in KMT2A is blown up out of scale to represent the mapped reads (blue) spanning over fusion breakpoint A at chr11:118357416 (+). Similarly, gene structure for *MLLT1* (NM_005934) along with preceding intergenic region is shown at the bottom with the reads (red) spanning over the intergenic fusion breakpoint B at chr19:6294605 (-). A middle part of the figure shows a structure of fusion contig carrying a portion of sequences from respective sides of breakpoints A (blue) and B (red) including insertion element (gray) in between. Representative portion of sequences of DNA from fusion partners and insertion element are shown accordingly.
*: Intergenic breakpoint B is not covered by SJPedPanel. Consequently, majority of the reference and fusion reads are sequenced from intronic breakpoint A, which is covered as part of *KMT2A* by the panel design.

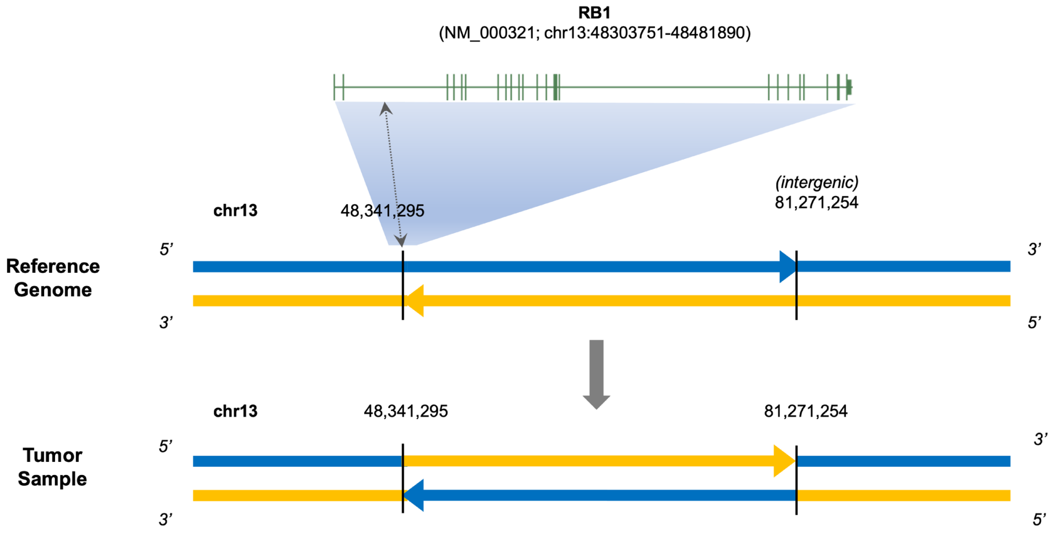


**Figure S11 – Complex genomic rearrangement missed using SJPedPanel.** An inversion involving RB1 gene in case SJRB0051 missed by SJPedPanel as the intergenic breakpoint is not covered by the panel. Only exonic regions are covered for RB1 gene in the panel.

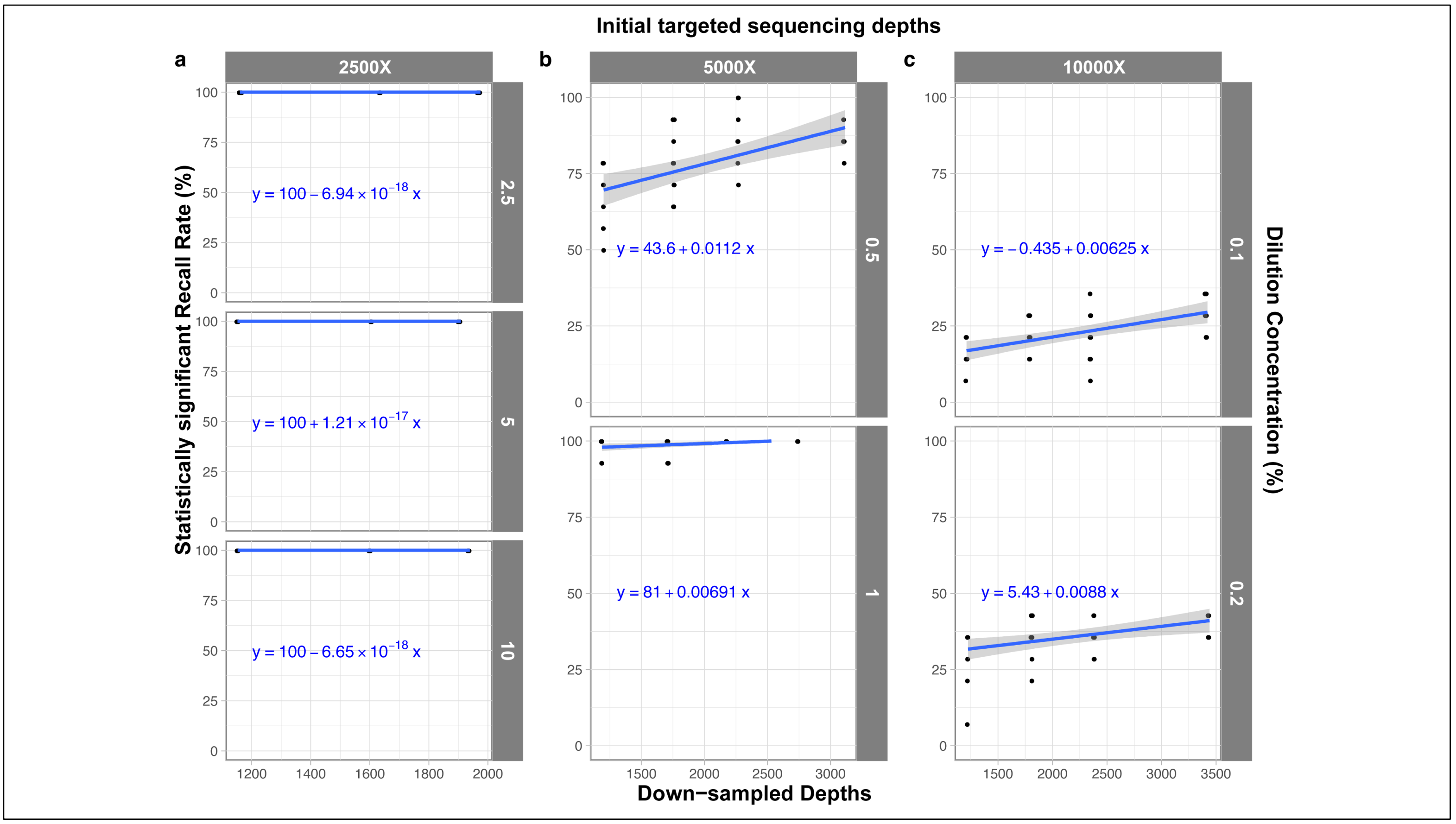


**Figure S12 – *In silico* down sampling experiment to determine optimal depth of sequencing and recall rate at various dilution concentrations.**

The X-axis shows down sampled depth and Y-axis shows recall rate using 14 SNVs with significant P-values ($P < 0.05$). (a) Down sampling results for samples originally sequenced at 2500X depth with 2.5% (top), 5% (middle) and 10% (bottom) dilution concentrations. (b) Down sampling results for samples originally sequenced at 5000X depth with 0.5% (top) and 1% (bottom) dilution concentrations. (c) Down sampling results for samples originally sequenced at 10,000X depth with 0.1% (top) and 0.2% (bottom) dilution concentrations.

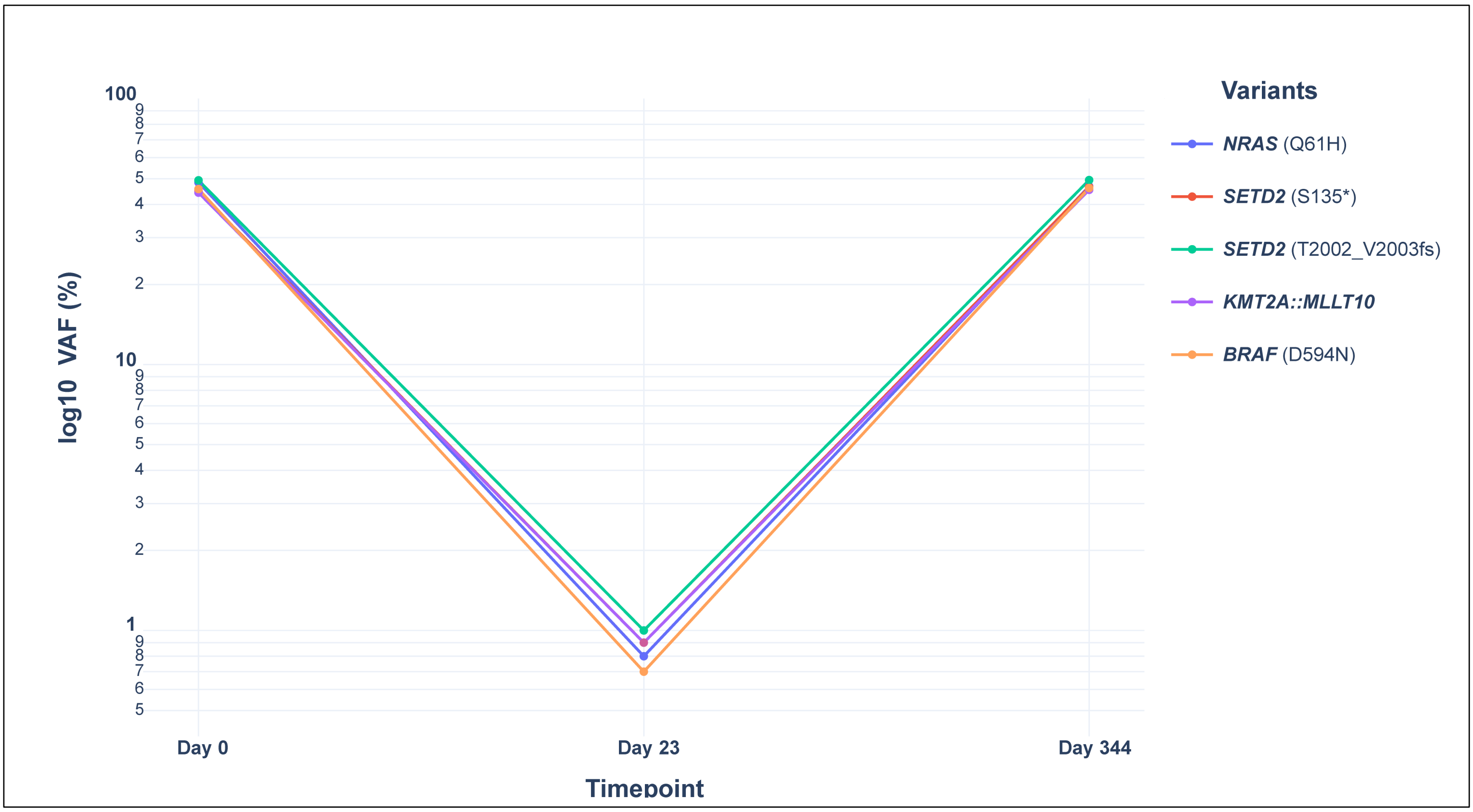


**Figure S13 – Real time tumor tracking in case SJAML016551.**

The X axis denotes three serial time points of sample collection from day of diagnosis (day 0), MRD (day 23) and relapse (day 344). Y-axis shows Variant Allele Frequency (%) on log10 scale. The variants are denoted in respective colored points/lines. See Additional file 1: Supplementary table ST24 for VAF% values.
