## Additional file 4 for "SJPedPanel: A pan-cancer gene panel for childhood malignancies"

Chromosomal distribution of SNPs (n=7590) in SJPedPanel

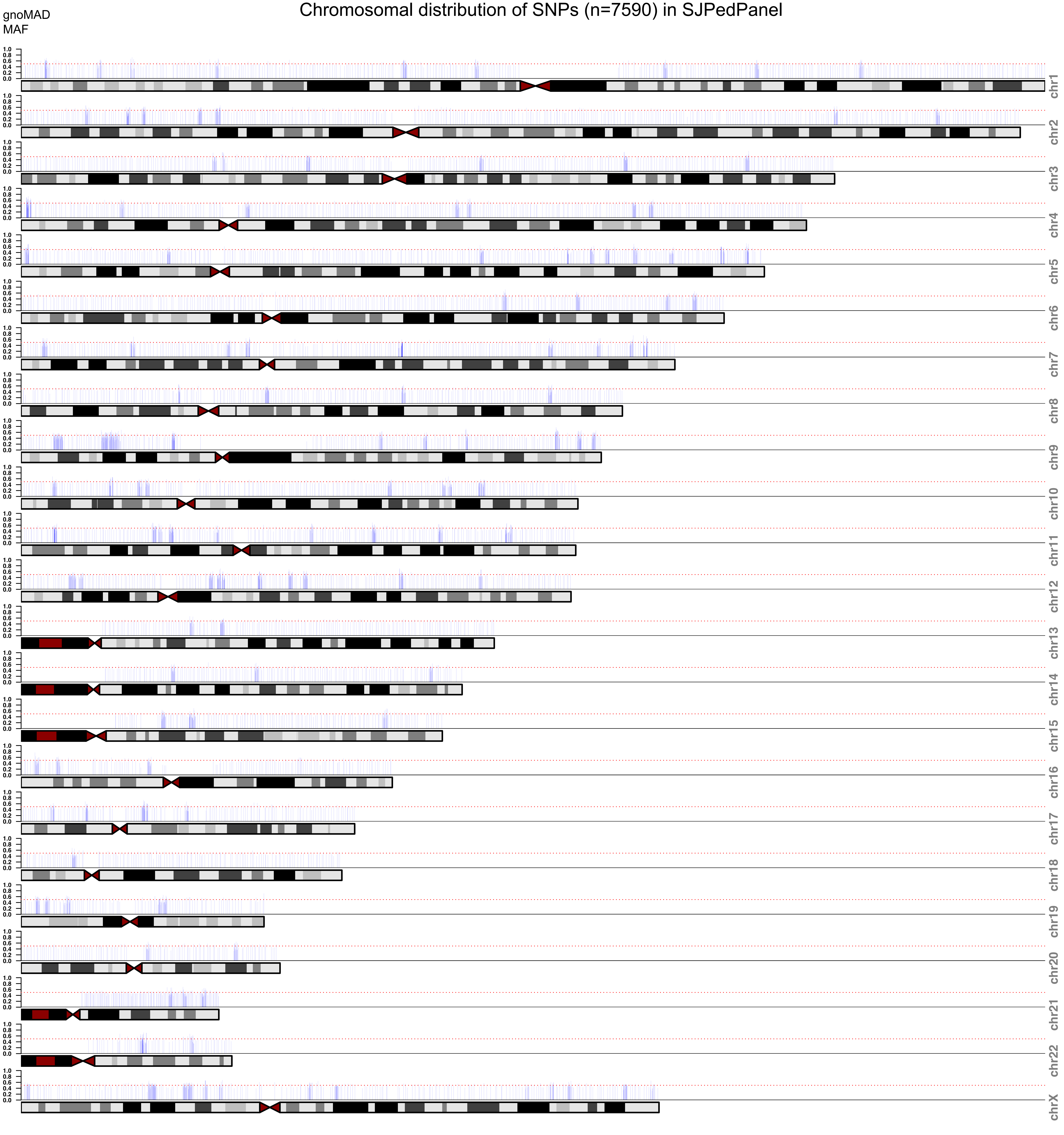
